# Exploratory analysis of immunization records highlights decreased SARS-CoV-2 rates in individuals with recent non-COVID-19 vaccinations

**DOI:** 10.1101/2020.07.27.20161976

**Authors:** Colin Pawlowski, Arjun Puranik, Hari Bandi, AJ Venkatakrishnan, Vineet Agarwal, Richard Kennedy, John C. O’Horo, Gregory J. Gores, Amy W. Williams, John Halamka, Andrew D. Badley, Venky Soundararajan

**Author notes:** Joint first authors (Colin Pawlowski, Arjun Puranik). Address correspondence to VS.

## Abstract

Multiple clinical studies are ongoing to assess whether existing vaccines may afford protection against SARS-CoV-2 infection through trained immunity. In this exploratory study, we analyze immunization records from 137,037 individuals who received SARS-CoV-2 PCR tests. We find that polio, Hemophilus influenzae type-B (HIB), measles-mumps-rubella (MMR), varicella, pneumococcal conjugate (PCV13), geriatric flu, and hepatitis A / hepatitis B (HepA-HepB) vaccines administered in the past 1, 2, and 5 years are associated with decreased SARS-CoV-2 infection rates, even after adjusting for geographic SARS-CoV-2 incidence and testing rates, demographics, comorbidities, and number of other vaccinations. Furthermore, age, race/ethnicity, and blood group stratified analyses reveal significantly lower SARS-CoV-2 rate among black individuals who have taken the PCV13 vaccine, with relative risk of 0.45 at the 5 year time horizon (n: 653, 95% CI: (0.32, 0.64), p-value: 6.9e-05). These findings suggest that additional pre-clinical and clinical studies are warranted to assess the protective effects of existing non-COVID-19 vaccines and explore underlying immunologic mechanisms. We note that the findings in this study are preliminary and are subject to change as more data becomes available and as further analysis is conducted.

## Introduction

Since the genome for SARS-CoV-2 was released on January 11, 2020, scientists around the world have been racing to develop a vaccine^1^. However, vaccine development is a long and expensive process, which takes on average over 10 years under ordinary circumstances^2^. Even for the previous epidemics of the past decade, including SARS, Zika, and Ebola, vaccines were not available before the virus spread was largely contained^3^.

Conventionally vaccinations are intended to train the adaptive immune system by generating an antigen-specific immune response. However, studies are also demonstrating that certain vaccines lead to protection against other infections through trained immunity^4^. For instance, vaccination against smallpox showed protection against measles and whooping cough^5^. Live vaccinia virus was successfully used against smallpox. Due to the urgent need to reduce the spread of COVID-19, scientists are turning to alternate methods to reduce the spread, such as repurposing existing vaccines. There are some hypotheses that the Bacillus Calmette–Guérin (BCG) and live poliovirus vaccines may provide some protective effect against SARS-CoV-2 infection^6,7^. There are several ongoing/recruiting clinical trials testing the protective effects of existing vaccines against SARS-CoV-2 infection, including: Polio^8^, Measles-Mumps-Rubella vaccine^9^, Influenza vaccine^10^, and BCG vaccine^11,12,13,14^.

In this work, we conduct a systematic analysis to determine whether or not a set of existing non-COVID-19 vaccines in the United States are associated with decreased rates of SARS-CoV-2 infection. In **Figure 1**, we provide an overview of the study design and statistical analyses. We consider data from 137,037 individuals from the Mayo Clinic electronic health record (EHR) database who received PCR tests for SARS-CoV-2 between February 15, 2020 and July 14, 2020 and have at least one ICD diagnostic code recorded in the past five years (see **Methods**). In **Table 1**, we show the clinical characteristics of the study population. In particular, 92,673 (67%) individuals have at least 1 vaccine in the past 5 years relative to the PCR testing date. In **Figure 2**, we present the SARS-CoV-2 infection rates for subsets of the study population with particular clinical covariates. We note that the rates of SARS-CoV-2 infection are higher in Black, Asian, and Hispanic racial and ethnic subgroups compared to the overall study population. This is likely due to higher rates of COVID-19 spread and/or decreased access to PCR testing. In addition, the rates of SARS-CoV-2 infection are lower in individuals with pre-existing conditions (e.g. hypertension, diabetes, obesity) possibly due to greater caution in avoiding exposure and/or higher PCR testing rates. Given this study population, we assess the rates of SARS-CoV-2 infection among individuals who did and did not receive one of 18 vaccines in the past 1, 2, and 5 years relative to the date of PCR testing. In **Table 2**, we present the full names, common formulations, and counts for the 18 vaccines that we consider.

**Figure 1:**
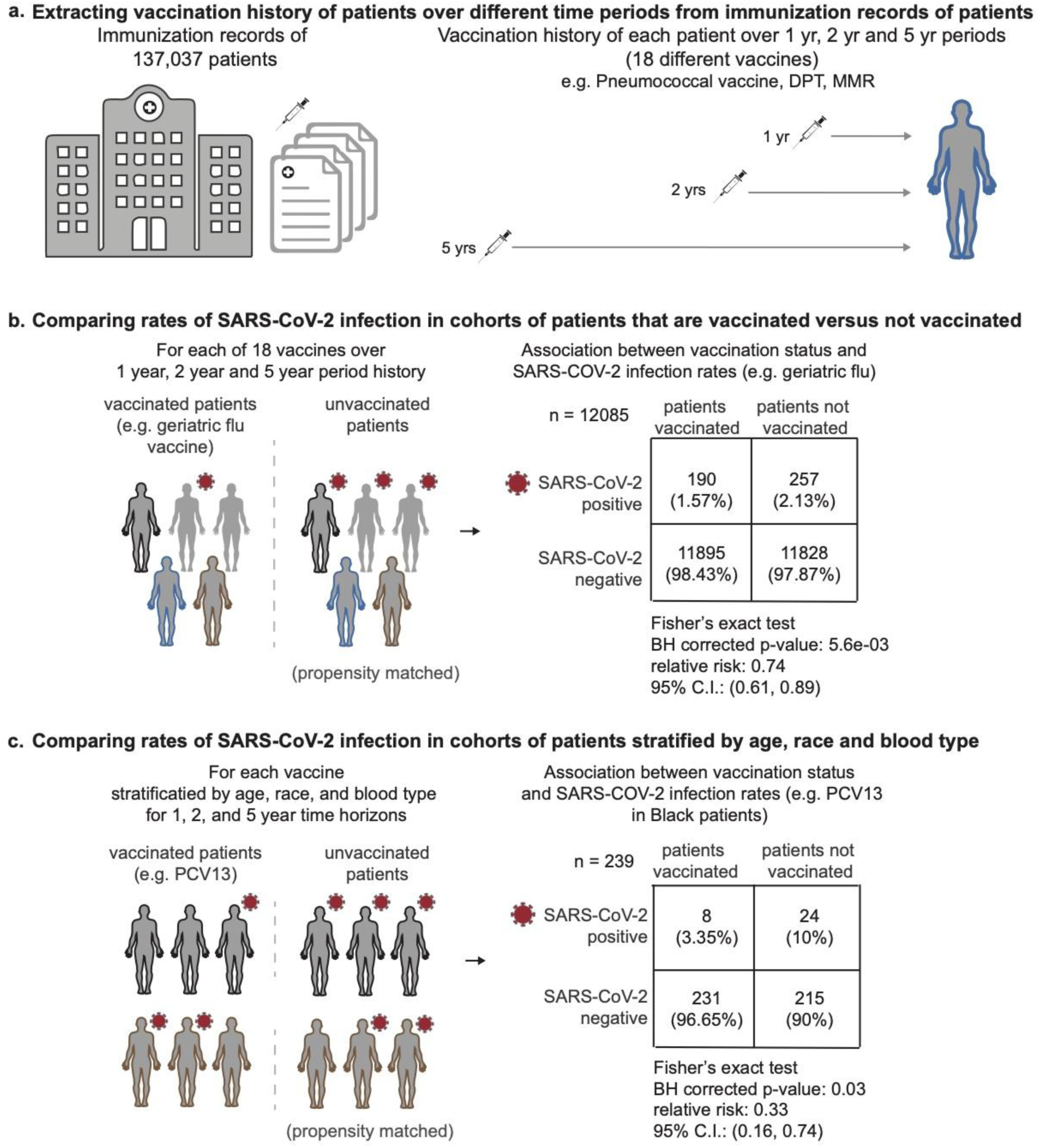
Overview of study design and statistical analyses. (A) Study design, datasets, and inclusion criteria used for the study; (B) Comparisons of SARS-CoV-2 rates between propensity-matched vaccinated and unvaccinated cohorts in the overall study population; (C) Comparisons of SARS-CoV-2 rates between propensity-matched vaccinated and unvaccinated cohorts in subgroups of the population stratified by age, race/ethnicity, and blood type.

**Table 1.**
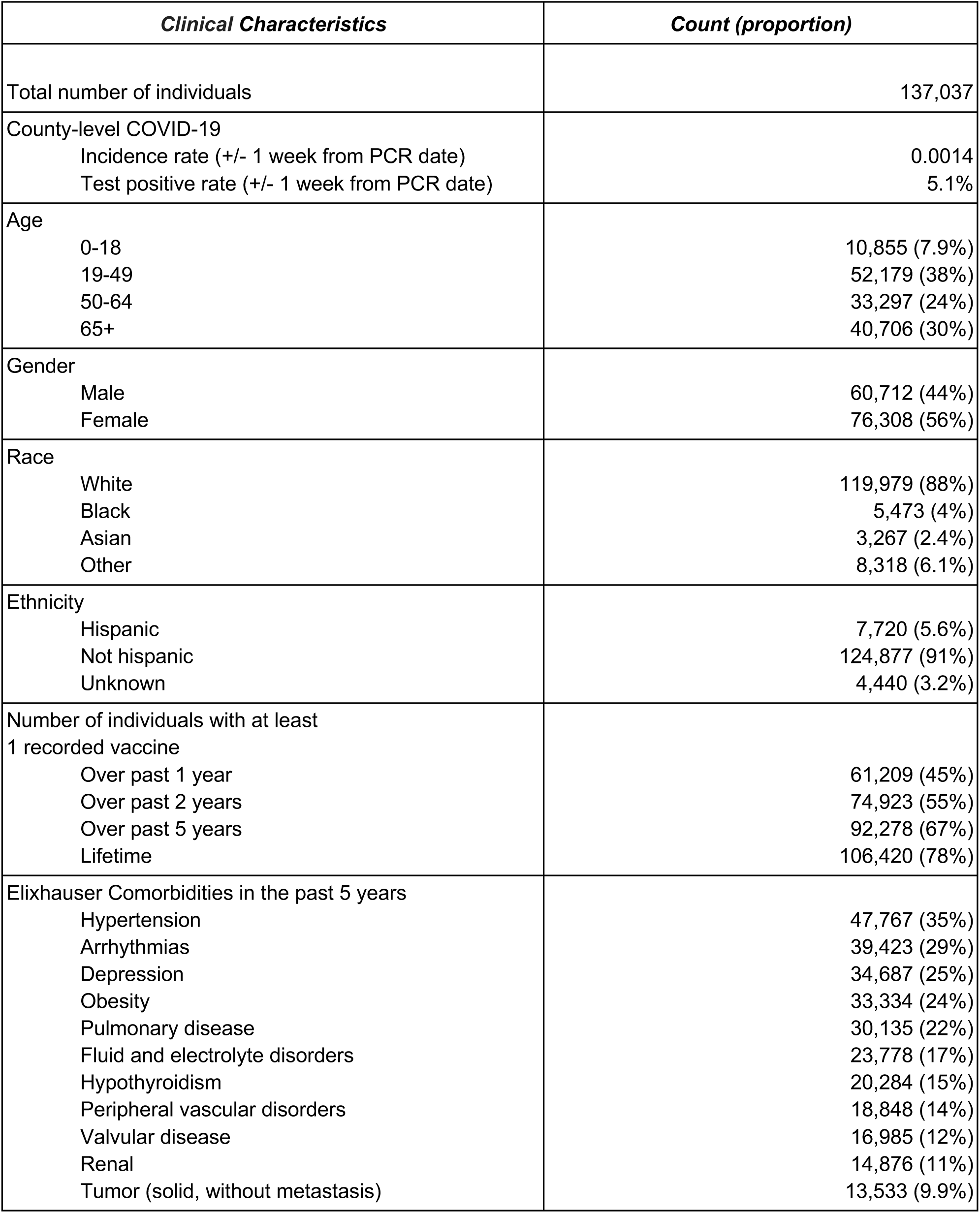
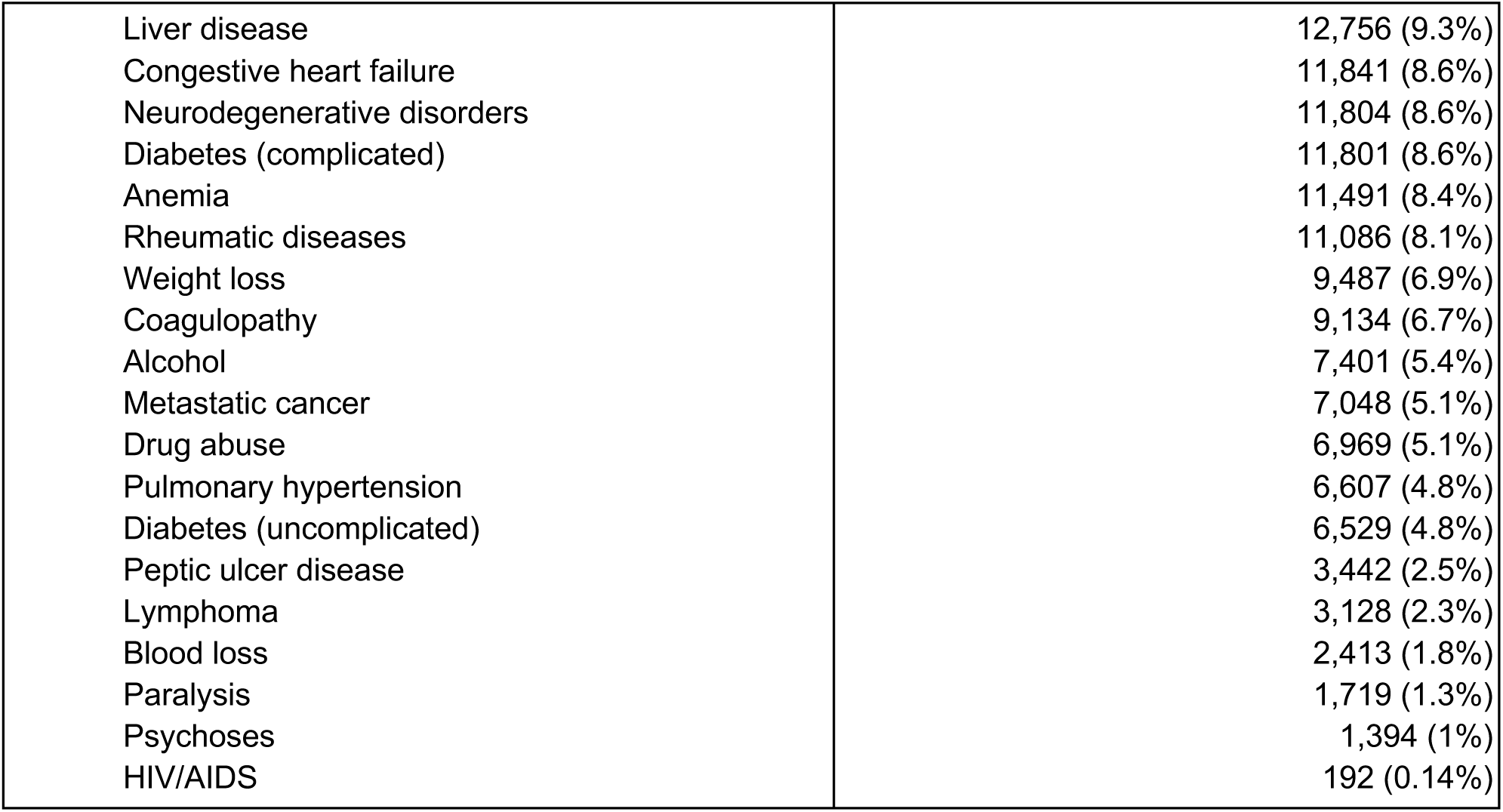
General characteristics of study population. Descriptive statistics for the study population, including: County-level COVID-19 incidence and testing rates, demographics (age, gender, race, ethnicity), vaccine counts, and Elixhauser comorbidities.

**Figure 2:**
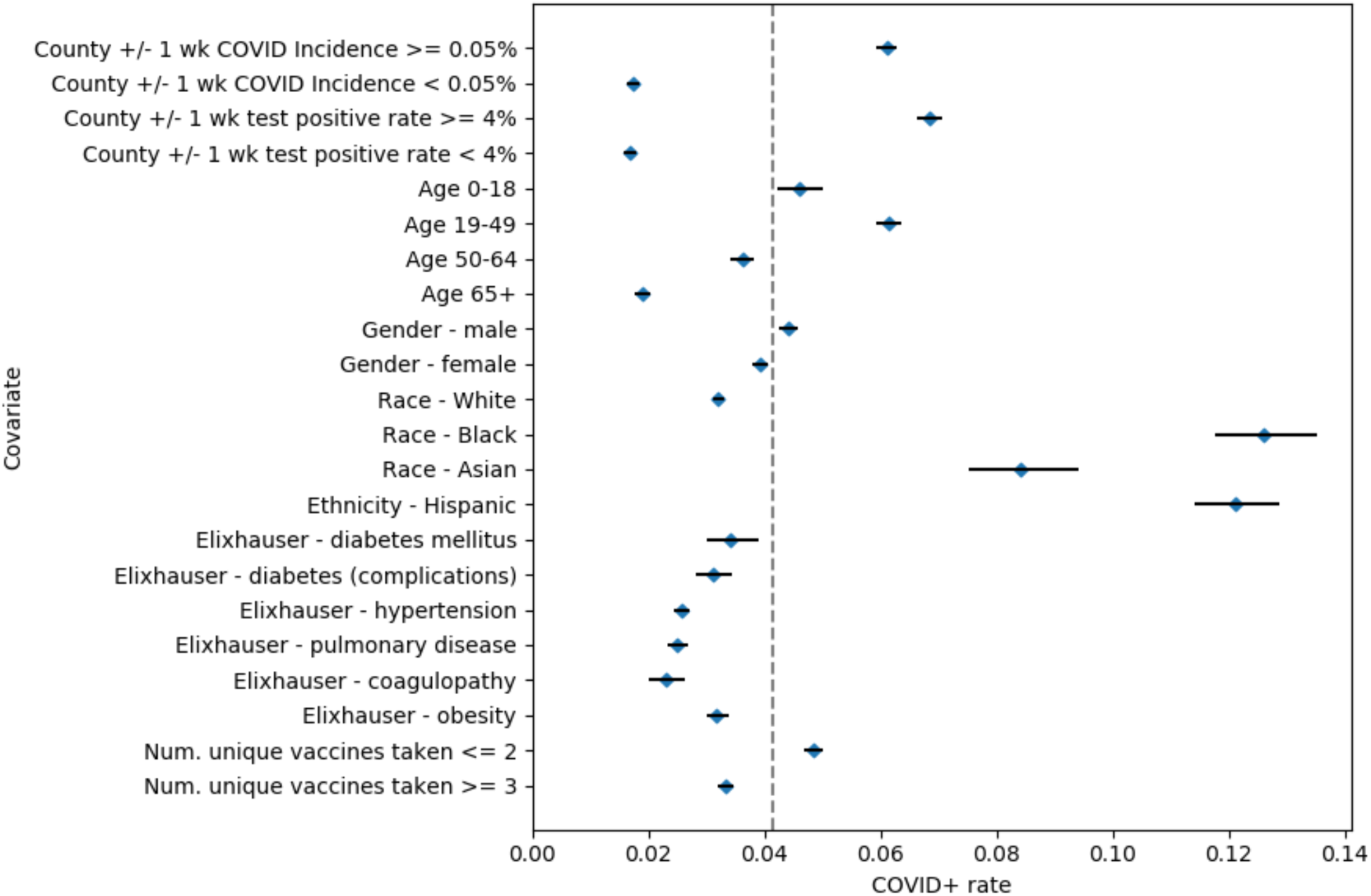
SARS-CoV-2 infection risk ratios by clinical covariate. SARS-CoV-2 rates among individuals with particular clinical covariates along with 95% confidence intervals. A dotted line indicates the study population SARS-CoV-2 rate of 4.4%. The clinical covariates include: county-level COVID-19 incidence and testing rates, age brackets (<18, 18-49, 50-64, 65+ years), gender, race, ethnicity, Elixhauser comorbidities, and number of unique vaccines taken.

**Table 2.** Summary of vaccines and common formulations. The 18 vaccines taken by at least 1,000 individuals within 5 years prior to their PCR test date, along with the most common formulations and patient counts. Note that some common formulations are combinations of multiple vaccines (e.g. Pentacel is a combination of DPT, polio, and HIB vaccines).

| **Vaccine name** | **Common formulations** | **Number of individuals taking in the past 5 years** |
| --- | --- | --- |
| Diphtheria-Pertussis-Tetanus (DPT) | TDAP; TD preservative free | 49,147 |
| Geriatric Flu | High dose geriatric (65+ years) | 22,290 |
| Haemophilus Influenzae type B (HIB) | DTAP-IPV/HIB (PENTACEL) | 4,651 |
| Human Papillomavirus (HPV) | 9VHPV, 4VHPV | 6,266 |
| Hepatitis A / Hepatitis B (HepA-HepB) | HepA adult; HepB adult<br>HepA pediatric/adolescent;<br>HepB pediatric/adolescent | 15,772 |
| Influenza (general) | Flublok/Fluarix/Fluzone | 78,043 |
| Influenza (live) | Influenza LAIV (Nasal) | 2,297 |
| Measles-mumps-rubella (MMR) | MMR, MMRV | 6,836 |
| Meningococcal | MCV4, MENB | 7,147 |
| Polio | DTAP-IPV/HIB (PENTACEL),<br>IPV, DTAP-IPV | 5,862 |
| Pediatric Flu | IIV4 | 11,676 |
| Pneumococcal conjugate (PCV13) | PCV13 | 25,954 |
| Pneumococcal polysaccharide (PPSV23) | PPSV23 | 17,422 |
| Rotavirus | RV5 (Rotateq) | 3,273 |
| RZV Zoster (Zostavax, Shingrix) | Zostavax, Shingrix | 17,630 |
| Tetanus | Td | 2,802 |
| Typhoid | TyVi, Ty21a | 2,393 |
| Varicella | VAR, MMRV | 5,783 |

First, we assess the overall association of vaccination status with the risk of SARS-CoV-2 infection (see **Methods**). We use propensity score matching to construct unvaccinated control groups for each of the vaccinated populations at the 1 year, 2 year, and 5-year time horizons. The unvaccinated control groups are balanced in covariates including demographics, county-level incidence and testing rates for SARS-CoV-2, comorbidities, and number of other vaccines taken in the past 5 years. Then, we compare the SARS-CoV-2 rates between each of the vaccinated cohorts and corresponding matched, unvaccinated control groups which have similar clinical characteristics. Second, we repeat the analysis on a set of age, race, and blood type stratified subgroups of the study population. In particular, for each subgroup, we run propensity score matching and compute the difference in SARS-CoV-2 infection rate between the vaccinated and unvaccinated (matched) cohorts. Finally, we run a series of sensitivity analyses to evaluate whether or not these results may be biased from unobserved confounders or other factors.

## Results

### Polio, HIB, MMR, Varicella, PCV13, Geriatric Flu, and HepA-HepB vaccines consistently show associations with lower rates of SARS-CoV-2 infection across 1, 2, and 5-year time horizons

The results of the propensity score matching for the 1 year, 2 year, and 5-year time horizons are presented in **Table 3, Table 4**, and **Table 5**, respectively. We observe that across all time horizons, Polio, Hemophilus Influenzae type B (HIB), Pneumococcal conjugate (PCV13), Geriatric Flu, Hepatitis A / Hepatitis B (HepA-HepB), and Measles-Mumps-Rubella (MMR) vaccinated cohorts show consistent lower rates of SARS-CoV-2 infection. In **Tables S1-S7**, we show the clinical characteristics for the vaccinated, unvaccinated, and matched cohorts for each of these vaccines at the 1-year time horizon. In **Figure 3**, we present the vaccination coverage rates for each of these vaccines in the study population for all time horizons.

**Table 3:**
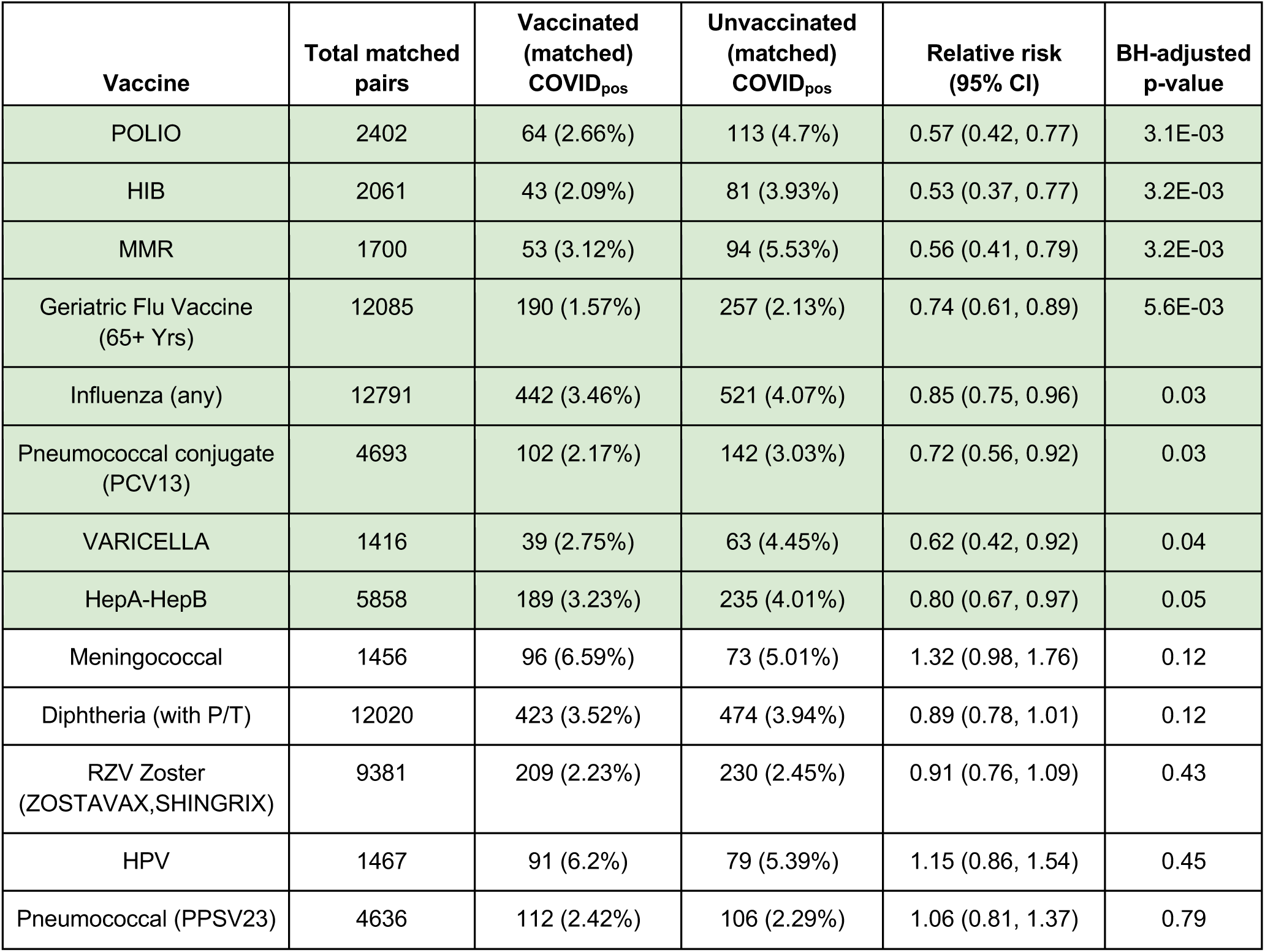
Summary of SARS-CoV-2 rates for vaccinated and unvaccinated propensity score matched cohorts (1 year time horizon). Table of SARS-CoV-2 infection rates for vaccinated and unvaccinated (matched) cohorts for vaccines administered within 1 year prior to PCR testing. Rows in which the SARS-CoV-2 rate is lower (adjusted p-value < 0.05) in the vaccinated cohort are highlighted in **green**, and rows in which the SARS-CoV-2 rate is lower in the unvaccinated cohort are highlighted in **orange**. The columns are **(1) Vaccine**: Name of the vaccine, **(2) Total matched pairs**: Number of pairs from the propensity matching procedure, which is the sample size of both vaccinated and unvaccinated cohorts after matching, **(3) Vaccinated (matched) COVID**_**pos**_: Number of COVID_pos_ cases among the vaccinated (matched) cohort, along with the percentage in parentheses, **(4) Unvaccinated (matched) COVID**_**pos**_: Number of COVID_pos_ cases among the unvaccinated (matched) cohort, along with the percentage in parentheses, **(5) Relative risk (95% CI):** Relative risk of COVID_pos_ in the vaccinated (matched) cohort compared to the unvaccinated (matched) cohort, along with 95% confidence interval in parentheses, **(6) BH-adjusted p-value:** Benjamini-Hochberg-adjusted Fisher exact test p-value.

| Vaccine | Total matched pairs | Vaccinated (matched) COVID<sub>pos</sub> | Unvaccinated (matched) COVID<sub>pos</sub> | Relative risk (95% CI) | BH-adjusted p-value |
| --- | --- | --- | --- | --- | --- |
| POLIO | 2402 | 64 (2.66%) | 113 (4.7%) | 0.57 (0.42, 0.77) | 3.1E-03 |
| HIB | 2061 | 43 (2.09%) | 81 (3.93%) | 0.53 (0.37, 0.77) | 3.2E-03 |
| MMR | 1700 | 53 (3.12%) | 94 (5.53%) | 0.56 (0.41, 0.79) | 3.2E-03 |
| Geriatric Flu Vaccine (65+ Yrs) | 12085 | 190 (1.57%) | 257 (2.13%) | 0.74 (0.61, 0.89) | 5.6E-03 |
| Influenza (any) | 12791 | 442 (3.46%) | 521 (4.07%) | 0.85 (0.75, 0.96) | 0.03 |
| Pneumococcal conjugate (PCV13) | 4693 | 102 (2.17%) | 142 (3.03%) | 0.72 (0.56, 0.92) | 0.03 |
| VARICELLA | 1416 | 39 (2.75%) | 63 (4.45%) | 0.62 (0.42, 0.92) | 0.04 |
| HepA-HepB | 5858 | 189 (3.23%) | 235 (4.01%) | 0.80 (0.67, 0.97) | 0.05 |
| Meningococcal | 1456 | 96 (6.59%) | 73 (5.01%) | 1.32 (0.98, 1.76) | 0.12 |
| Diphtheria (with P/T) | 12020 | 423 (3.52%) | 474 (3.94%) | 0.89 (0.78, 1.01) | 0.12 |
| RZV Zoster (ZOSTAVAX, SHINGRIX) | 9381 | 209 (2.23%) | 230 (2.45%) | 0.91 (0.76, 1.09) | 0.43 |
| HPV | 1467 | 91 (6.2%) | 79 (5.39%) | 1.15 (0.86, 1.54) | 0.45 |
| Pneumococcal (PPSV23) | 4636 | 112 (2.42%) | 106 (2.29%) | 1.06 (0.81, 1.37) | 0.79 |

**Table 4:** Summary of SARS-CoV-2 rates for vaccinated and unvaccinated propensity score matched cohorts (2 year time horizon). Table of SARS-CoV-2 infection rates for vaccinated and unvaccinated (matched) cohorts for vaccines administered within 2 years prior to PCR testing. Rows in which the SARS-CoV-2 rate is lower (adjusted p-value < 0.05) in the vaccinated cohort are highlighted in **green**, and rows in which the SARS-CoV-2 rate is lower in the unvaccinated cohort are highlighted in **orange**. The columns are **(1) Vaccine**: Name of the vaccine, **(2) Total matched pairs**: Number of pairs from the propensity matching procedure, which is the sample size of both vaccinated and unvaccinated cohorts after matching, **(3) Vaccinated (matched) COVID**_**pos**_: Number of COVID_pos_ cases among the vaccinated (matched) cohort, along with the percentage in parentheses, **(4) Unvaccinated (matched) COVID**_**pos**_: Number of COVID_pos_ cases among the unvaccinated (matched) cohort, along with the percentage in parentheses, **(5) Relative risk (95% CI):** Relative risk of COVID_pos_ in the vaccinated (matched) cohort compared to the unvaccinated (matched) cohort, along with 95% confidence interval in parentheses, **(6) BH-adjusted p-value:** Benjamini-Hochberg-adjusted Fisher exact test p-value.

| Vaccine | Total matched pairs | Vaccinated (matched) COVID<sub>pos</sub> | Unvaccinated (matched) COVID<sub>pos</sub> | Relative risk (95% CI) | BH-adjusted p-value |
| --- | --- | --- | --- | --- | --- |
| POLIO | 2821 | 88 (3.12%) | 173 (6.13%) | 0.51 (0.40, 0.66) | 1.2E-06 |
| HepA-HepB | 8443 | 265 (3.14%) | 379 (4.49%) | 0.70 (0.60, 0.82) | 4.0E-05 |
| Pneumococcal conjugate (PCV13) | 8285 | 185 (2.23%) | 275 (3.32%) | 0.67 (0.56, 0.81) | 9.5E-05 |
| HIB | 2711 | 56 (2.07%) | 110 (4.06%) | 0.51 (0.37, 0.70) | 9.5E-05 |
| Meningococcal | 3009 | 223 (7.41%) | 155 (5.15%) | 1.44 (1.18, 1.75) | 1.1E-03 |
| TYPHOID | 1051 | 72 (6.85%) | 36 (3.43%) | 2.00 (1.35, 2.93) | 1.2E-03 |
| VARICELLA | 2544 | 85 (3.34%) | 135 (5.31%) | 0.63 (0.48, 0.82) | 1.5E-03 |
| MMR | 3055 | 111 (3.63%) | 161 (5.27%) | 0.69 (0.55, 0.87) | 4.3E-03 |
| RZV Zoster (ZOSTAVAX, SHINGRIX) | 14000 | 290 (2.07%) | 360 (2.57%) | 0.81 (0.69, 0.94) | 0.01 |
| Pneumococcal (PPSV23) | 8751 | 210 (2.4%) | 265 (3.03%) | 0.79 (0.66, 0.95) | 0.02 |
| Geriatric Flu Vaccine (65+ Yrs) | 12360 | 217 (1.76%) | 269 (2.18%) | 0.81 (0.68, 0.96) | 0.03 |
| Diphtheria (with P/T) | 21705 | 805 (3.71%) | 879 (4.05%) | 0.92 (0.83, 1.01) | 0.09 |
| Influenza (any) | 17652 | 665 (3.77%) | 723 (4.1%) | 0.92 (0.83, 1.02) | 0.14 |
| HPV | 2634 | 177 (6.72%) | 168 (6.38%) | 1.05 (0.86, 1.29) | 0.70 |
| ROTAVIRUS | 612 | 10 (1.63%) | 8 (1.31%) | 1.25 (0.50, 3.03) | 0.81 |

**Table 5:** Summary of SARS-CoV-2 rates for vaccinated and unvaccinated propensity score matched cohorts (5 year time horizon). Table of SARS-CoV-2 infection rates for vaccinated and unvaccinated (matched) cohorts for vaccines administered within 5 years prior to PCR testing. Rows in which the SARS-CoV-2 rate is lower (adjusted p-value < 0.05) in the vaccinated cohort are highlighted in **green**, and rows in which the SARS-CoV-2 rate is lower in the unvaccinated cohort are highlighted in **orange**. The columns are **(1) Vaccine**: Name of the vaccine, **(2) Total matched pairs**: Number of pairs from the propensity matching procedure, which is the sample size of both vaccinated and unvaccinated cohorts after matching, **(3) Vaccinated (matched) COVID**_**pos**_: Number of COVID_pos_ cases among the vaccinated (matched) cohort, along with the percentage in parentheses, **(4) Unvaccinated (matched) COVID**_**pos**_: Number of COVID_pos_ cases among the unvaccinated (matched) cohort, along with the percentage in parentheses, **(5) Relative risk (95% CI):** Relative risk of COVID_pos_ in the vaccinated (matched) cohort compared to the unvaccinated (matched) cohort, along with 95% confidence interval in parentheses, **(6) BH-adjusted p-value:** Benjamini-Hochberg-adjusted Fisher exact test p-value.

| Vaccine | Total matched pairs | Vaccinated (matched) COVID<sub>pos</sub> | Unvaccinated (matched) COVID<sub>pos</sub> | Relative risk (95% CI) | BH-adjusted p-value |
| --- | --- | --- | --- | --- | --- |
| Pneumococcal conjugate (PCV13) | 23194 | 503 (2.17%) | 745 (3.21%) | 0.68 (0.60, 0.76) | 7.2E-11 |
| Meningococcal | 7008 | 552 (7.88%) | 366 (5.22%) | 1.51 (1.33, 1.71) | 2.1E-09 |
| POLIO | 3072 | 131 (4.26%) | 213 (6.93%) | 0.62 (0.50, 0.76) | 3.8E-05 |
| HPV | 6179 | 494 (7.99%) | 376 (6.09%) | 1.31 (1.15, 1.49) | 1.7E-04 |
| Geriatric Flu Vaccine (65+ Yrs) | 13860 | 246 (1.77%) | 330 (2.38%) | 0.75 (0.63, 0.88) | 1.7E-03 |
| HIB | 2913 | 73 (2.51%) | 120 (4.12%) | 0.61 (0.46, 0.81) | 2.2E-03 |
| MMR | 4965 | 226 (4.55%) | 299 (6.02%) | 0.76 (0.64, 0.89) | 3.1E-03 |
| RZV Zoster (ZOSTAVAX, SHINGRIX) | 16889 | 355 (2.1%) | 440 (2.61%) | 0.81 (0.70, 0.93) | 5.7E-03 |
| TYPHOID | 2383 | 149 (6.25%) | 103 (4.32%) | 1.45 (1.13, 1.84) | 7.0E-03 |
| HepA-HepB | 13377 | 541 (4.04%) | 628 (4.69%) | 0.86 (0.77, 0.96) | 0.02 |
| VARICELLA | 3623 | 175 (4.83%) | 218 (6.02%) | 0.80 (0.66, 0.97) | 0.05 |
| Pediatric Flu Vaccine | 11676 | 426 (3.65%) | 375 (3.21%) | 1.14 (0.99, 1.30) | 0.11 |
| TETANUS | 2800 | 88 (3.14%) | 67 (2.39%) | 1.31 (0.96, 1.79) | 0.14 |
| Influenza (Live) | 2296 | 110 (4.79%) | 87 (3.79%) | 1.26 (0.96, 1.66) | 0.14 |
| Diphtheria (with P/T) | 40334 | 1590 (3.94%) | 1678 (4.16%) | 0.95 (0.89, 1.01) | 0.14 |
| Pneumococcal (PPSV23) | 16836 | 414 (2.46%) | 446 (2.65%) | 0.93 (0.81, 1.06) | 0.32 |
| Influenza (any) | 22057 | 985 (4.47%) | 955 (4.33%) | 1.03 (0.95, 1.13) | 0.53 |
| ROTAVIRUS | 694 | 18 (2.59%) | 23 (3.31%) | 0.78 (0.43, 1.43) | 0.53 |

**Figure 3:**
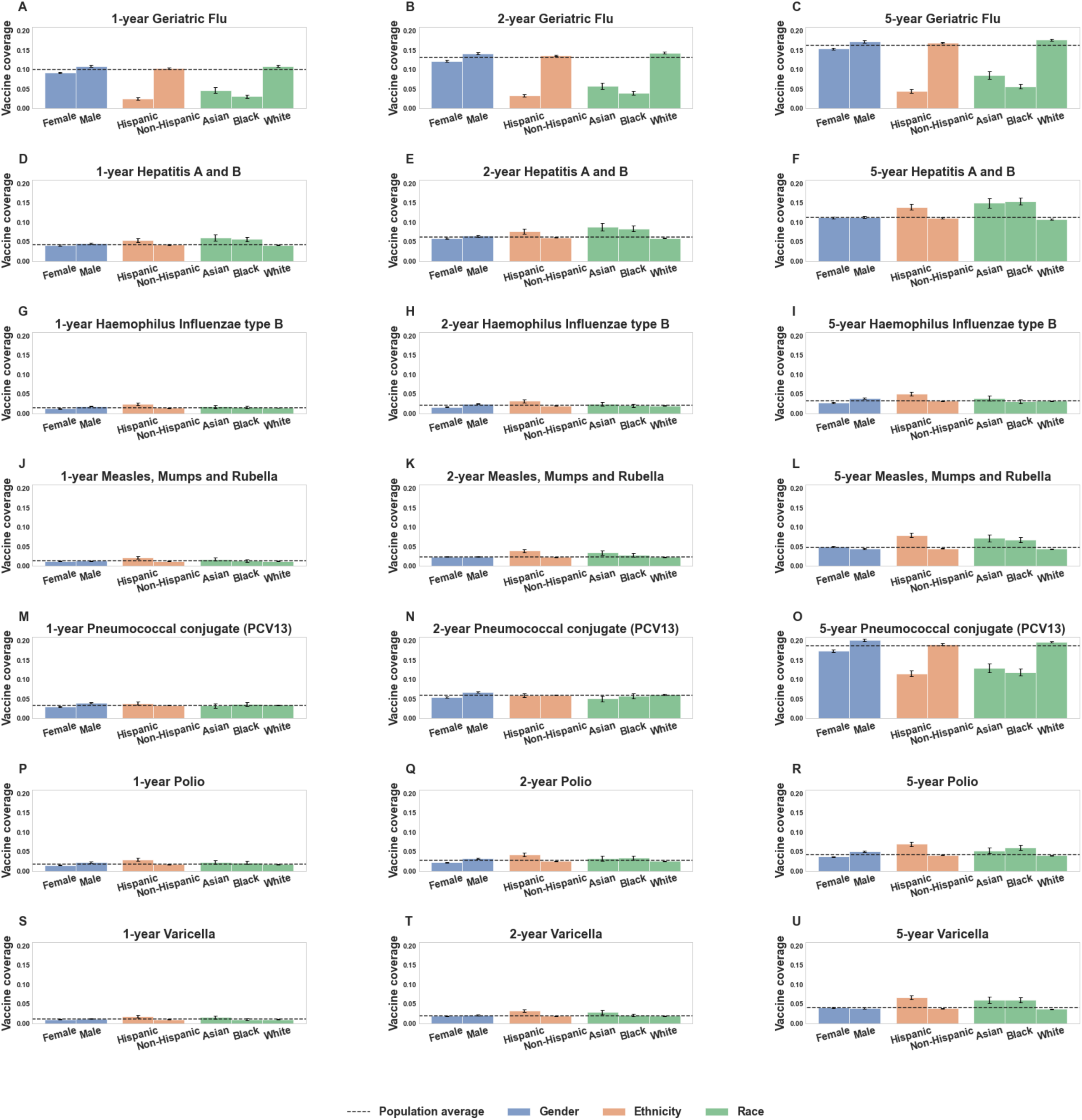
Vaccination coverage plots. Coverage rates for vaccines associated with lower SARS-CoV-2 rates, stratified by different demographic factors (age, race/ethnicity, and gender), along with 95% confidence intervals. In each plot, the population average vaccination rate for the (vaccine, time horizon) pair is shown as a horizontal line. Includes coverage rates for the following vaccines for the past 1, 2, and 5 year time horizons: **(A-C)** Geriatric Flu vaccine, **(D-F)** Hepatitis A / Hepatitis B (HepA-HepB), **(G-I)** Haemophilus Influenzae type B (HIB), **(J-L)** Measles-Mumps-Rubella (MMR), **(M-O)** Pneumococcal Conjugate (PCV13), **(P-R)** Polio, **(S-U)** Varicella.

Overall, we observe that the Polio and HIB vaccinated cohorts generally have the lowest relative risks for SARS-CoV-2 infection across all time horizons. The relative risk of SARS-CoV-2 infection is 0.57 (n: 2,402, 95% CI: (0.42, 0.77), p-value: 0.003) for individuals who have taken the Polio vaccine in the past 1 year, and 0.53 (n: 2,061, (95% CI: (0.37, 0.77), p-value: 3.2e-03) for individuals who have taken the HIB vaccine in the past year. We note that these vaccines are almost exclusively administered to individuals under 18 years of age, as shown in **Figure 4**. Other vaccines that are commonly administered to younger individuals with strong negative correlations with SARS-CoV-2 infection include MMR and Varicella vaccines.

**Figure 4:**
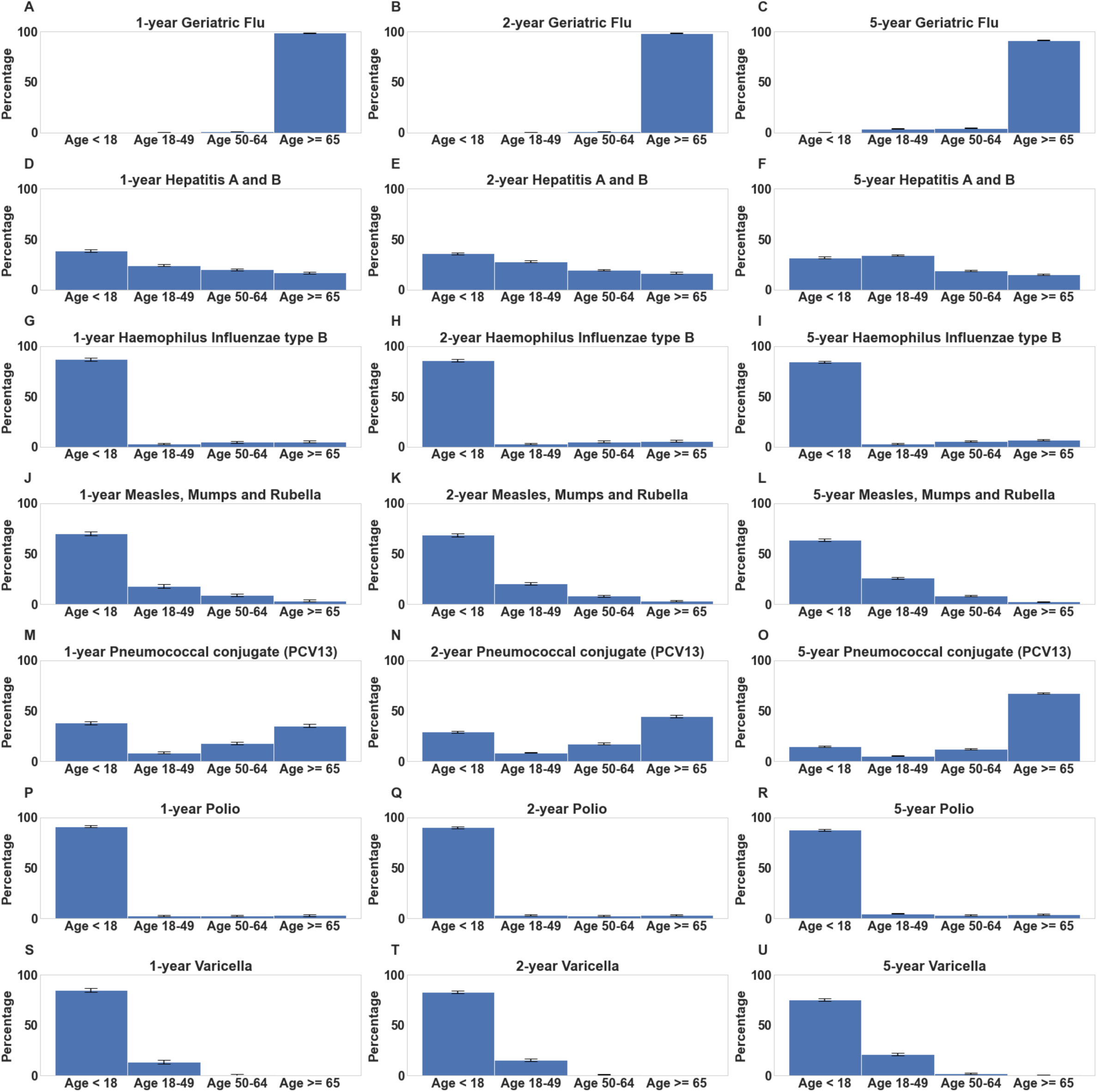
Age distribution plots for vaccinated cohorts. Distributions of age in cohorts of individuals who received vaccines associated with lower SARS-CoV-2 rates. For each vaccine, percentages of vaccinated individuals within age brackets (<18, 18-49, 50-64, 65+ years) are shown along with 95% confidence intervals. Includes age distributions for the following vaccines for the past 1, 2, and 5 year time horizons: **(A-C)** Geriatric Flu vaccine, **(D-F)** Hepatitis A / Hepatitis B (HepA-HepB), **(G-I)** Haemophilus Influenzae type B (HIB), **(J-L)** Measles-Mumps-Rubella (MMR), **(M-O)** Pneumococcal Conjugate (PCV13), **(P-R)** Polio, **(S-U)** Varicella.

The other vaccines which are consistently associated with lower SARS-CoV-2 rates include PCV13, Geriatric Flu, and HepA-HepB vaccines. At the 1 year time horizon, the relative risks of SARS-CoV-2 infection are 0.72 for PCV13 (n: 4,693, 95% CI: (0.56, 0.92), p-value: 0.03), 0.74 for Geriatric Flu (n: 12,085, 95% CI: (0.61, 0.89), p-value: 5.6e-03), and 0.80 for HepA-HepB (n: 5,858, 95% CI: (0.67, 0.97), p-value: 0.05). Although the relative risks are less significant compared to Polio and HIB, these associations may be particularly interesting to explore further because these vaccines are commonly administered across a broader age range of the population (see **Figure 4**).

### Pairwise correlation analysis reveals strong associations between administration of HIB, Polio, Rotavirus, Varicella, and MMR vaccines

In order to identify vaccines which may be confounding factors for other vaccines that are linked to reduced rates of SARS-CoV-2 infection, we conduct a pairwise correlation analysis. For example, it is possible that the lower rates of SARS-CoV-2 infection that we observe for one vaccine are in fact caused by another vaccine which is highly correlated with the former. To measure the correlations we use Cohen’s kappa, which is a measure of correlation for categorical variables that ranges from −1 to +1. In particular, Cohen’s kappa = +1 indicates that the pair of vaccines are always administered together, Cohen’s kappa = 0 indicates that the pair of vaccines are independent of each other, and Cohen’s kappa = −1 indicates that the pair of vaccines are never administered together.

In **Figure 5**, we present a heatmap of the pairwise correlations for each of the 18 vaccines administered in the 5 years prior to the PCR test date. Sorted by Cohen’s kappa value, the top vaccine pairs with kappa ≥ 0.60 are: HIB and Rotavirus (0.83), HIB and Polio (0.80), MMR and Varicella (0.74), Polio and Varicella (0.72), Polio and Rotavirus (0.71), MMR and Polio (0.68). From this, we see that there is a cluster of vaccines which are commonly administered together, which includes: HIB, Polio, Rotavirus, Varicella, and MMR vaccines. The majority of individuals who receive this cluster of vaccines are children <18 years old (see **Figure 4**). We note that in this cluster, the vaccines HIB, Polio, Varicella, and MMR are all consistently associated with lower SARS-CoV-2 rates. This suggests that some of the lower rates of SARS-CoV-2 observed in these vaccinated cohorts may be confounded by the other vaccines in this group.

**Figure 5:**
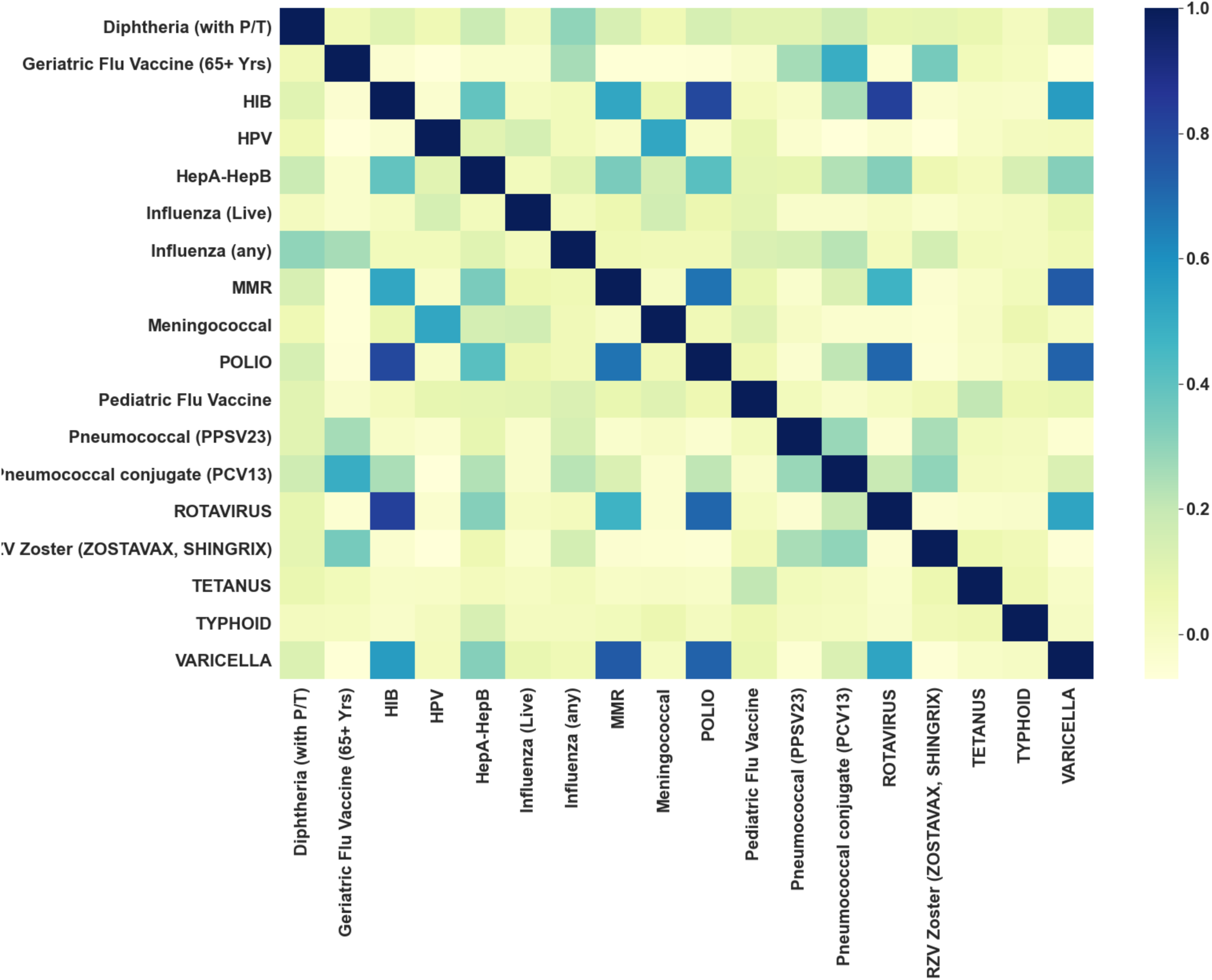
Heatmap of pairwise vaccine correlations. Heatmap showing correlations between pairs of vaccines based upon their administration to the same patient within the past 5 years. Each cell in this plot is shaded according to its Cohen’s kappa value, a measure of correlation for categorical variables that ranges from −1 to +1. Cohen’s kappa = +1 indicates that the pair of vaccines are always administered together, Cohen’s kappa = 0 indicates that the pair of vaccines are independent of each other, and Cohen’s kappa = −1 indicates that the pair of vaccines are never administered together.

### Stratification by race reveals that Polio, HIB, and PCV13 vaccines are associated with lower SARS-CoV-2 rates in particular racial subgroups across 1, 2, and 5-year time periods

In **Tables 7-9**, we present the results of propensity score matching at the 1, 2, and 5-year time horizon, respectively, on study cohorts stratified by race. We observe that PCV13 vaccination is linked with significantly decreased SARS-CoV-2 rates in the Black subpopulation. In particular, the relative risk of SARS-CoV-2 infection for black individuals who have been administered PCV13 is 0.24 at the 1 year time horizon (n: 197, 95% CI: (0.09, 0.71), p-value: 0.03); 0.33 at the 2 year time horizon (n: 239, 95% CI: (0.16, 0.74), p-value: 0.03); and 0.45 at the 5 year time horizon (n: 653, 95% CI (0.32, 0.64), p-value: 6.9e-5). Furthermore, at the 5-year time horizon, the relative risk for the PCV13 vaccinated cohort of black individuals is significantly lower than the relative risk for the PCV13 vaccinated cohort overall (p-value: 0.03).

In addition, we observe that Polio, HIB, and PCV13 vaccines are linked with decreased SARS-CoV-2 rates in the White subpopulation. However, since 119,979 (88%) of individuals in the study population are white, the relative risks for these vaccinated cohorts are close to the relative risks for the overall population (see **Tables 3-5**). Matching within subgroups was done by age group (0-18, 19-49, 50-64, 65+) and blood group (A, B, AB, O) as well, but no significant within-subgroup associations between any vaccine and SARS-CoV-2 rates were found. This suggests that associations between vaccines and SARS-CoV-2 infection rates may not be strongly specific to particular age ranges/blood groups.

### Sensitivity Analysis

#### Tipping point analysis shows that associations between reduced SARS-CoV-2 rates and Polio vaccine (1, 2 year time horizons), PCV13 (5 year time horizon) are most robust to unobserved confounders

In this retrospective study, we evaluate the correlations between vaccination and SARS-CoV-2 infection, taking into account a number of possible confounding variables, such as demographic variables and geographic COVID-19 incidence rate (see **Methods**). However, it is possible that the results from this study have been influenced by unobserved confounders. For example, we do not explicitly control for travel history, which was a significant risk factor for SARS-CoV-2 infection early on in the pandemic.

In **Figure 6**, we present the results from the tipping point analysis on the statistically significant associations between vaccination and reduced rates of SARS-CoV-2 infection in the overall study population. For each time horizon, we show the relative prevalence and effect size that would be required for an unobserved confounder to overturn the conclusion for a given (vaccine, time horizon) pair. For reference, we show the effect size of the covariate (county-level COVID-19 incidence rate ≥ median value) as a potential confounder, which has a large relative risk of 2.78.

**Figure 6:**
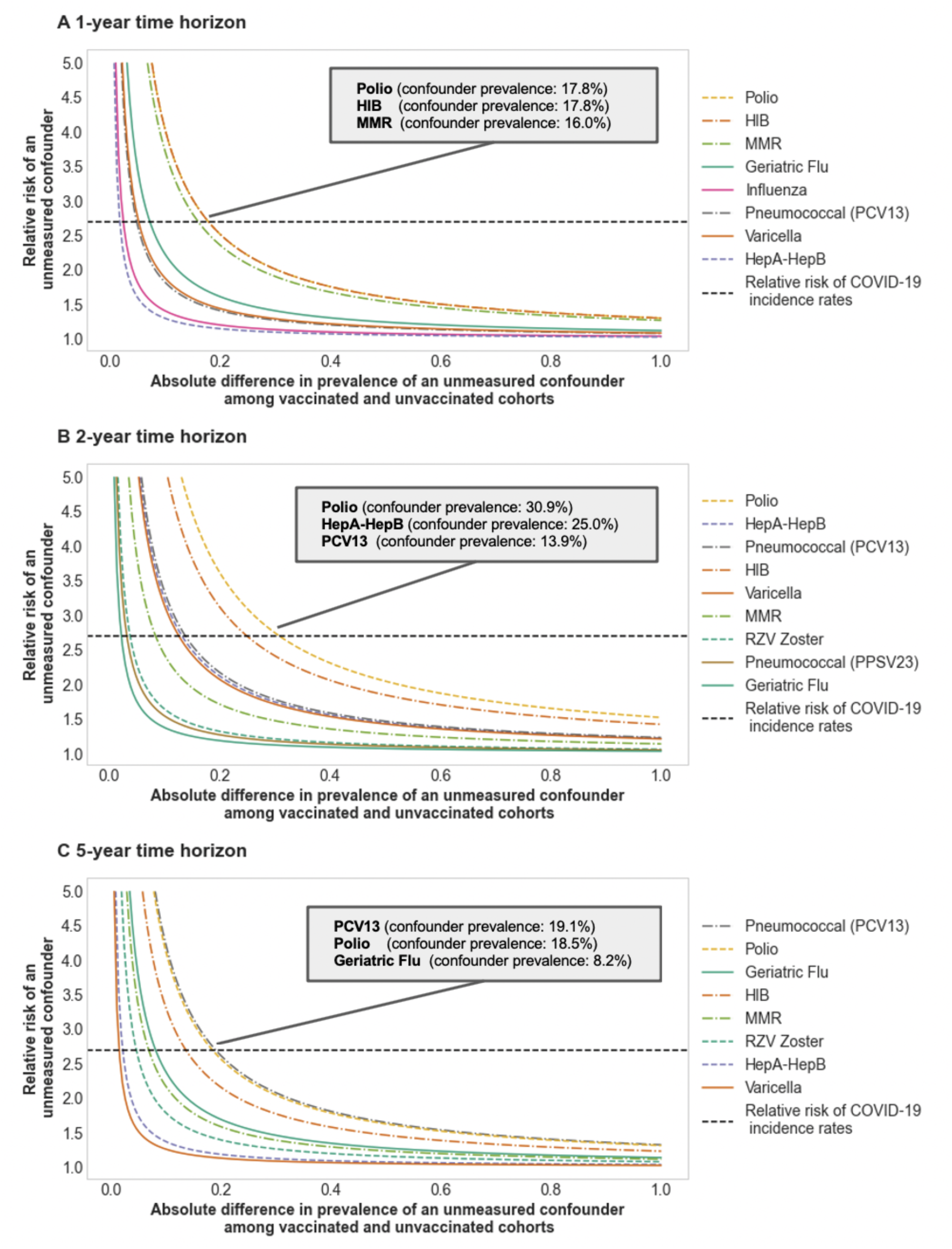
Sensitivity of associations between vaccines and SARS-CoV-2 rates to unobserved confounders. Tipping point analysis for associations of vaccines and lower rates of SARS-CoV-2 infection for **(A)** 1-year, **(B)** 2-year, and **(C)** 5-year time horizons. For each vaccine that is associated with lower SARS-CoV-2 rates in a particular time horizon, we plot the (prevalence, effect size) combinations of an unobserved confounder that would be required to overturn the results. The x-axis indicates the absolute difference in prevalence of the confounder between vaccinated and unvaccinated (matched) cohorts. For example, if the unobserved confounder is present in 25% of the vaccinated cohort and 5% of the unvaccinated cohort, then the absolute difference in prevalence would be 20%. The y-axis indicates the relative COVID_pos_ risk (effect size) of the unobserved confounder. For reference, we show the relative risk of (county-level COVID-19 incidence rate ≥ median value) as a horizontal dotted line, which is equal to 2.78. Each plot is annotated with the top 3 vaccines that are most robust to unobserved confounders, along with the intersection point between the vaccine curve and the reference line. For example, for the polio vaccine at the 1 year time horizon, an unobserved confounder with a relative risk of 2.78 which is prevalent in 17.8% of the vaccinated cohort and 0% of the unvaccinated cohort could explain the differences in SARS-CoV-2 infection rates that we observe in the data.

At the 1 year and 2 year time horizons, the associations of the Polio vaccine to lower rates of SARS-CoV-2 infection are most robust to the impact of a potential unobserved confounder. In particular, an unobserved confounder with a large effect size of 2.78 would need to have an absolute difference in prevalence between vaccinated and unvaccinated cohorts of 17.8% (30.9%) in order to overturn the results for the 1 year (2 year) time horizon. On the other hand, at the 5 year time horizon, the association of PCV13 and lower rates of SARS-CoV-2 infection is most robust to the influence by unobserved confounders. An unobserved confounder with a large effect size of 2.78 would need to have an absolute difference in prevalence between vaccinated and unvaccinated cohorts of 19.1% in order to render the findings insignificant.

## Discussion

Ongoing clinical studies offer preliminary evidence that existing vaccines may reduce risk of SARS-CoV-2 infection. For example, interim results from the ACTIVATE trial^12^ indicate that the BCG vaccine reduces SARS-CoV-2 infection rates up to 53%. While specific vaccines such as BCG are being tested for cross-protective effects against SARS-CoV-2 infection based upon their prior potential for protection against other diseases^14^, to our knowledge, a systematic hypothesis-free analysis to identify potential vaccines that can have beneficial effects against SARS-CoV-2 infection is lacking. Our retrospective study has analyzed 18 different vaccines and identified key vaccines that are correlated with lower rates of SARS-CoV-2 infection after controlling for confounding factors (see **Results**). In particular, we find that individuals who have been recently vaccinated with one of Polio, HIB, MMR, Varicella, PCV13, Geriatric Flu, or HepA-HepB vaccines have lower rates of SARS-CoV-2 infection. These vaccines are promising candidates for follow-up pre-clinical animal studies and clinical trials. We note that this list of vaccines is preliminary and may change as more data becomes available and as further analysis is conducted.

For the rest of the 18 vaccines that we considered, the correlations with SARS-CoV-2 infection were either insignificant or varied across the time horizons of interest. In some cases, these vaccines may serve as negative controls in clinical trials testing the safety and efficacy of novel COVID-19 vaccines. For example, a clinical trial evaluating the COVID-19 vaccine candidate ChAdOx1 uses Meningococcal vaccine as a comparator arm^15^. Preliminary results from this trial indicate that as expected, Meningococcal vaccine does not induce antibody responses against SARS-CoV-2 spike protein. It may be interesting to evaluate the antibody responses for some of the vaccines that we have found to be significantly correlated with lower rates of SARS-CoV-2 infection, to explore if there is any underlying immunologic mechanism for the associations that we observe.

Because the BCG vaccine is rarely administered in the US, this vaccine did not meet the sample size threshold for inclusion in our analysis. From the limited data available, there were 51 individuals in the study population who had taken BCG vaccine in the past 5 years, and among these 0 individuals tested positive for SARS-CoV-2 infection (95% CI: (0.0%, 7.0%)). Among the 198 individuals who had taken BCG vaccine at least once in their lifetime, there were 6 (3.0%) individuals who tested positive for SARS-CoV-2 infection (95% CI: (1.4%, 6.5%)). As a result, more data from additional medical centers would be required for us to assess the associations between BCG vaccine and SARS-CoV-2 infection.

Due to the observational nature of this study, there are potential biases which may have impacted the findings, including confounding, selection bias, and measurement bias. The motivation for using propensity score matching was to account for confounding. Although we take into account some potential confounders through propensity score matching, there may still be residual confounding from unobserved factors (e.g. socioeconomic status, indications, contraindications, etc.) which may be different for each vaccine. For example, travel history is a risk factor for exposure to SARS-CoV-2 infection that we do not explicitly account for in this study. Our motivation for the tipping point sensitivity analysis is to estimate the effect size and prevalence of an unobserved confounder which would be required to overturn the statistically significant findings (see **Figure 6**). Even among the variables that we consider, there is potential for bias if the cohorts are poorly matched on those covariates. In **Tables S1-S7**, we present the propensity score matching results for a number of vaccines at the 1 year time horizon, in order to show the matching quality for each of these statistical comparisons. Furthermore, we present plots showing the distribution of the age covariate in particular in **Figure S1**. We note that for some vaccines, differences in age between the vaccinated and unvaccinated (matched) cohorts may have influenced the results.

In addition, it is possible that restricting the study population to SARS-CoV-2 PCR tested individuals may have introduced selection bias. For example, vaccinated individuals may engage in more health-seeking behaviors to reduce their potential COVID-19 risk, and also have a higher likelihood of seeking out a PCR test. This type of bias is known as the “healthy user effect”, which is suspected to have influenced the findings of recent COVID-19 observational studies^16,17^. We performed sensitivity analyses using breast cancer and colon cancer screening as negative controls which suggest that the propensity score matching analysis is in part effective in filtering out healthy user effect for the associations between vaccination status and SARS-CoV-2 risk. Finally, measurement bias is a concern as vaccination records may be incomplete for some individuals in our cohort since they may have received the vaccines outside of the Mayo Clinic system. We plan to perform additional sensitivity analyses to further explore these potential sources of bias.

As an initial exploratory analysis linking historical vaccination records to SARS-CoV-2 PCR testing results, more research is warranted in order to confirm the findings. We plan to update this analysis in coming months as more PCR testing data becomes available. Also, we note that this study is based on data from one academic medical center in the United States, which restricts the analysis to vaccines administered in this geographic region. Notably, we do not have sufficient immunization record data on the BCG vaccine, which has shown promise in early clinical trials. As a result, the findings from this study would be well complemented by similar studies from hospitals across the world.

## Methods

### Study design

This is an observational study in a cohort of individuals who underwent polymerase chain reaction (PCR) testing for suspected SARS-CoV-2 infection at the Mayo Clinic and hospitals affiliated to the Mayo health system. The full dataset includes 152,548 individuals who received PCR tests between February 15, 2020 and July 14, 2020. We restricted the study population to 137,037 individuals from this dataset who have at least one ICD code recorded in the past 5 years. This exclusion criteria is applied in order to restrict the analysis to individuals with medical history data. Within this PCR tested cohort, we define COVID_pos_ to be persons with at least one positive PCR test result for SARS-CoV-2 infection, which includes 5,679 individuals. Similarly, we define COVID_neg_ to be persons with all negative PCR test results, which includes 131,358 individuals.

For the study population of 137,037 individuals, we obtain a number of clinical covariates from the Mayo Clinic electronic health record (EHR) database, including: demographics (age, gender, race, ethnicity, county of residence), ICD diagnostic billing codes from the past 5 years, and immunization records from the past 5 years (68 unique vaccines; we focus on the 18 taken by at least 1,000 individuals over the past 5 years). We use the Elixhauser Comorbidity Index to map the ICD codes from each individual from the past 5 years to a set of 30 medically relevant comorbidities^18^. In addition to the Mayo Clinic EHR database, we use the Corona Data Scraper online database to obtain incidence rates of COVID-19 at the county-level in the United States^18,19^. By linking the county of residence data from the EHR with the incidence rates of COVID-19 from Corona Data Scraper, we are able to obtain county-level incidence rates of COVID-19 for 136,313 individuals in the study population. We also obtain county-level testing data for 100,433 individuals in the study population from (i) Minnesota state government records and (ii) public county-level testing data scraped from other state/county websites. In **Table 1**, we present the average values for each of the clinical covariates in the study population.

Given these clinical covariates, we conduct a series of statistical analyses to assess whether or not each of the 19 vaccines has an association with lower rates of SARS-CoV-2 infection at the 1 year, 2 years, and 5 year time horizons. For each vaccine and time horizon, the vaccinated cohort is defined as the set of individuals in the study population who received the vaccine within the past time horizon. For example, the “2-year polio vaccinated cohort” is the set of individuals who received the polio vaccine within the past two years. Similarly, for each vaccine and time horizon, the unvaccinated cohort is defined as the set of individuals in the study population who did not receive the vaccine within the past time horizon. For example, the “5-year influenza unvaccinated cohort” is the set of individuals who did not receive the influenza vaccine within the past five years.

In the following sections, we describe the statistical methods that we use to compare the rates of COVID-19 between the vaccinated and unvaccinated cohorts for each of the (vaccine, time horizon) pairs. First, we describe the propensity score matching analysis to construct unvaccinated control groups that have similar clinical characteristics to the vaccinated cohorts. Second, we describe the statistical tests that we use to determine which of the (vaccine, time horizon) pairs have the most significant association with lower rates of SARS-CoV-2 infection for the 1 year, 2 year, and 5 year time horizons, both overall and for particular demographic subgroups. Third, we describe the covariate-level stratification analysis to identify vaccines which have the largest association with lower rates of SARS-CoV-2 infection for particular demographic subgroups. Finally, we describe the sensitivity analyses that we use to evaluate the robustness of the statistical methods to potential biases from unobserved confounders or other factors that could impact the overall results from this observational study.

### Propensity score matching to construct unvaccinated control groups

Before running the propensity score matching step, first we filtered to vaccinated cohorts with at least 1,000 persons. For the overall statistical analysis, there were 13, 15, and 18 vaccines which met this threshold for the 1 year, 2 year, and 5 year time horizons, respectively.

For each vaccinated cohort with sufficient numbers of individuals, we applied 1:1 propensity score matching to construct a corresponding unvaccinated control group with similar clinical characteristics^20^. We refer to this as the “unvaccinated (matched)” cohort, which is a subset of the unvaccinated cohort. We considered the following clinical covariates in the propensity score matching step:

- **Demographics** (Age, Gender, Race, Ethnicity)
- **County-level COVID-19 incidence rate:** (Number of positive SARS-CoV-2 PCR tests in county) / (Total population of county) within +/-1 week of PCR testing date.
- **County-level COVID-19 test positive rate:** (Number of positive SARS-CoV-2 PCR tests in county) / (Number of PCR tests in county) within +/-1 week of PCR testing date.
- **Elixhauser comorbidities:** Medical history derived from ICD diagnostic billing codes in the past 5 years relative to the PCR testing date. Includes indicators for the following conditions: (1) congestive heart failure, (2) cardiac arrhythmias, (3) valvular disease, (4) pulmonary circulation disorders, (5) peripheral vascular disorders, (6) hypertension, (7) paralysis, (8) neurodegenerative disorders, (9) chronic pulmonary disease, (10) diabetes, (11) diabetes with complications, (12) hypothyroidism, (13) renal failure, (14) liver disease, (15) peptic ulcer disease (excluding bleeding), (16) AIDS/HIV, (17) lymphoma, (18) metastatic cancer, (19) solid tumor without metastasis, (20) rheumatoid arthritis/collagen vascular diseases, (21) coagulopathy, (22) obesity, (23) weight loss, (24) fluid and electrolyte disorders, (25) blood loss anemia, (26) deficiency anemia, (27) alcohol abuse, (28) drug abuse, (29) psychoses, (30) depression.
- **Pregnancy:** Whether or not the individual had a pregnancy-related ICD code recorded in the past 90 days relative to the PCR testing date.
- **Number of other vaccines:** Count of the total number of unique vaccines (excluding the vaccine which is the treatment variable) taken by the individual in the past 5 years relative to the PCR testing date.

For each of the vaccinated cohorts, we fit a logistic regression model to predict whether or not the individual was vaccinated, using these covariates as predictors. We trained the logistic regression model using the `scikit-learn` package in Python^21^. Then, we used the model-predicted probability of an individual receiving the vaccine as the propensity score for the individual. Matching was done without replacement using greedy nearest-neighbor matching within calipers. Some subjects were dropped from the positive cohort in this procedure. The matching was performed with caliper width 0.2 * (pooled standard deviation of scores), as suggested in the literature^22^.

### Statistical assessment of associations between vaccines and SARS-CoV-2 infection rates for the overall study population

After the propensity score matching step, we compare the COVID_pos_ rates for the vaccinated and unvaccinated (matched) cohorts. First, we compute the relative risk, which is equal to the COVID_pos_ rate for the vaccinated (matched) cohort divided by the COVID_pos_ rate for the unvaccinated (matched) cohort. We use a Fisher exact test to compute the p-value for this association. We then apply the Benjamini-Hochberg (BH) adjustment^23^ on the p-values over all vaccines for each time horizon to control the False Discovery Rate (at 0.05). We also compute and report 95% confidence intervals for the relative risks.

### Statistical assessment of associations between vaccines and SARS-CoV-2 infection rates for age, race/ethnicity, and blood type stratified subgroups

We repeat the statistical analysis on subsets of the study population stratified by age, race/ethnicity, and blood type. For age, we consider the subgroups: 0 to 18 years, 19 to 49 years old, 50 to 64 years old, and ≥ 65 years old. For race/ethnicity, we consider the subgroups: White, Black, Asian, and Hispanic. For blood type, we consider the subgroups: O, A, B, and AB. We note that age and race/ethnicity were recorded in the dataset for all subjects, but blood type information was only available for 41,828 subjects.

For each vaccine, at the 1, 2, and 5 year time horizons, we use propensity score matching to construct unvaccinated control groups for each age bracket, race/ethnicity, and blood type subgroup. Matching was done on the same covariates as in the overall analysis (apart from the Race/Ethnicity covariates for the race/ethnicity subgroups). We then compared the COVID_pos_ rates between the vaccinated and unvaccinated (matched) cohorts, and reported the relative risk, 95% confidence interval, and BH-corrected p-values.

### Sensitivity Analyses

We performed two sets of sensitivity analyses, as described below.

### Cancer screens as negative controls for propensity score matching procedure

To assess the effectiveness of the propensity score matching procedure, we ran the statistical analysis using cancer screens as the exposure variable instead of vaccinations (i.e. negative control exposure). This set of experiments serves as a negative control because it is highly unlikely that cancer screenings are causally linked to risk of SARS-CoV-2 infection. In particular, we considered the following two cancer screens as negative controls:

- **Colon cancer screen:** Whether or not the individual received a screening for colon cancer (within a specified time horizon relative to PCR testing date).
- **Mammogram:** Whether or not the individual received a mammogram screening for breast cancer (within a specified time horizon relative to PCR testing date),

In **Table 6**, we present the results from the negative control experiments. In the unmatched cohorts, we observe that persons who have had a mammogram in the past 1, 2, or 5 years have significantly lower rates of SARS-CoV-2 infection compared to persons who have not had mammograms during the same time period. For example, the SARS-CoV-2 infection rate is 2.5% among persons with mammograms in the past 5 years and 4.5% among persons without mammograms in the past 5 years (p-value: 1.9e-47). This significant difference in SARS-CoV-2 infection rate can be explained by confounding variables, because the unmatched cohorts have different underlying clinical characteristics. However, after propensity score matching, the SARS-CoV-2 infection rate is 2.8% among persons with mammograms in the past 5 years and 2.8% among persons without mammograms in the past 5 years (p-value: 1).

**Table 6:**
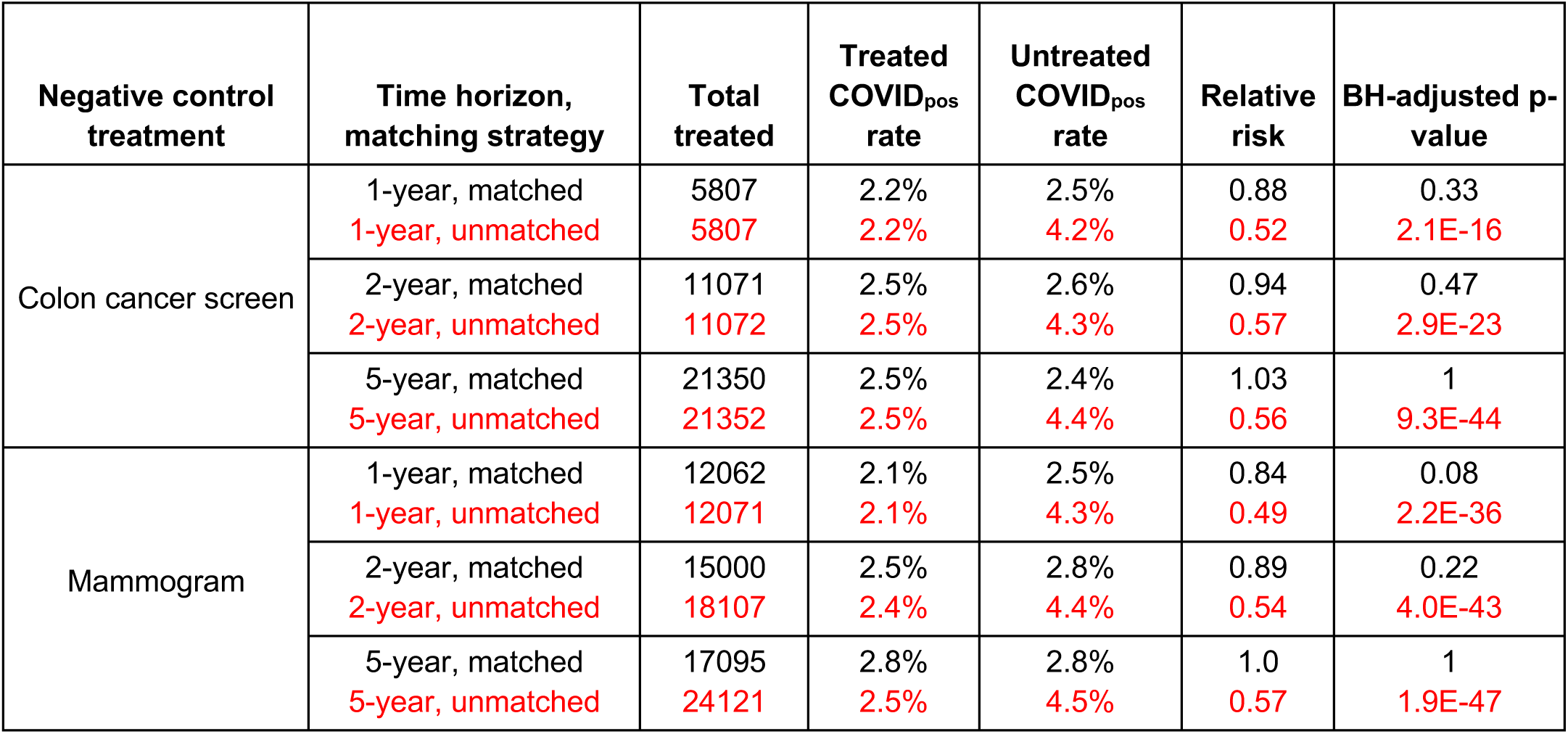
Summary of SARS-CoV-2 rates for individuals who did vs. did not receive negative control treatments before and after propensity score matching. SARS-CoV-2 positive rates, relative risks, and associated BH-adjusted Fisher exact p-values for individuals who received or did not receive negative control treatments over the past 1 year, 2 years, and 5 years prior to PCR test. The negative control treatments considered are: (1) Colon cancer screen and (2) Mammogram. The BH adjustment is applied per time horizon, as in the main analysis. Numbers are shown before and after propensity score matching. Unmatched numbers are shown in red text.

| Negative control treatment | Time horizon, matching strategy | Total treated | Treated COVID<sub>pos</sub> rate | Untreated COVID<sub>pos</sub> rate | Relative risk | BH-adjusted p-value |
| --- | --- | --- | --- | --- | --- | --- |
| Colon cancer screen | 1-year, matched<br>1-year, unmatched | 5807<br>5807 | 2.2%<br>2.2% | 2.5%<br>4.2% | 0.88<br>0.52 | 0.33<br>2.1E-16 |
|  | 2-year, matched<br>2-year, unmatched | 11071<br>11072 | 2.5%<br>2.5% | 2.6%<br>4.3% | 0.94<br>0.57 | 0.47<br>2.9E-23 |
|  | 5-year, matched<br>5-year, unmatched | 21350<br>21352 | 2.5%<br>2.5% | 2.4%<br>4.4% | 1.03<br>0.56 | 1<br>9.3E-44 |
| Mammogram | 1-year, matched<br>1-year, unmatched | 12062<br>12071 | 2.1%<br>2.1% | 2.5%<br>4.3% | 0.84<br>0.49 | 0.08<br>2.2E-36 |
|  | 2-year, matched<br>2-year, unmatched | 15000<br>18107 | 2.5%<br>2.4% | 2.8%<br>4.4% | 0.89<br>0.54 | 0.22<br>4.0E-43 |
|  | 5-year, matched<br>5-year, unmatched | 17095<br>24121 | 2.8%<br>2.5% | 2.8%<br>4.5% | 1.0<br>0.57 | 1<br>1.9E-47 |

**Table 7:**
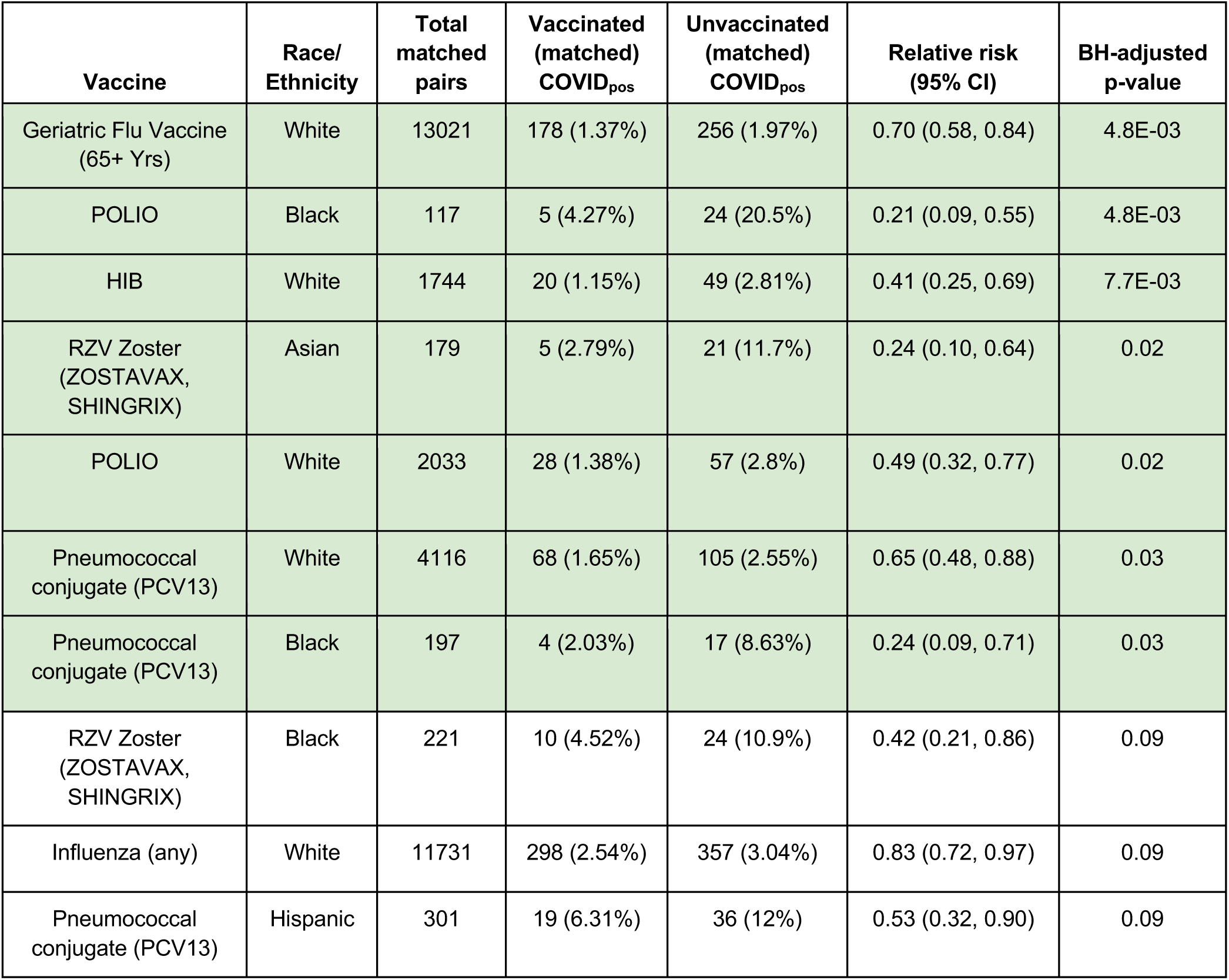
Summary of SARS-CoV-2 rates for race/ethnicity-stratified vaccinated and unvaccinated propensity score matched cohorts (1 year time horizon). Table of SARS-CoV-2 infection rates for vaccinated and unvaccinated (matched) race/ethnicity subgroup cohorts for vaccines administered within 1 year prior to PCR testing. Only rows with adjusted p-values ≤ 0.1 are included. Rows in which the SARS-CoV-2 rate is lower (adjusted p-value < 0.05) in the vaccinated cohort are highlighted in **green**, and rows in which the SARS-CoV-2 rate is lower in the unvaccinated cohort are highlighted in **orange**. The columns are **(1) Vaccine**: Name of the vaccine, **(2) Race/ethnicity**: Race/ethnicity subgroup, **(3) Total matched pairs**: Number of pairs from the propensity matching procedure, which is the sample size of both vaccinated and unvaccinated cohorts after matching, **(4) Vaccinated (matched) COVID**_**pos**_: Number of COVID_pos_ cases among the vaccinated (matched) cohort, along with the percentage in parentheses, **(5) Unvaccinated (matched) COVID**_**pos**_: Number of COVID_pos_ cases among the unvaccinated (matched) cohort, along with the percentage in parentheses, **(6) Relative risk (95% CI):** Relative risk of COVID_pos_ in the vaccinated (matched) cohort compared to the unvaccinated (matched) cohort, along with 95% confidence interval in parentheses, **(7) BH-adjusted p-value:** Benjamini-Hochberg-adjusted Fisher exact test p-value.

**Table 8:** Summary of SARS-CoV-2 rates for race/ethnicity-stratified vaccinated and unvaccinated propensity score matched cohorts (2 year time horizon). Table of SARS-CoV-2 infection rates for vaccinated and unvaccinated (matched) race/ethnicity subgroup cohorts for vaccines administered within 5 years prior to PCR testing. Only rows with adjusted p-values ≤ 0.1 are included. Rows in which the SARS-CoV-2 rate is lower (adjusted p-value < 0.05) in the vaccinated cohort are highlighted in **green**, and rows in which the SARS-CoV-2 rate is lower in the unvaccinated cohort are highlighted in **orange**. The columns are **(1) Vaccine**: Name of the vaccine, **(2) Race/ethnicity**: Race/ethnicity subgroup, **(3) Total matched pairs**: Number of pairs from the propensity matching procedure, which is the sample size of both vaccinated and unvaccinated cohorts after matching, **(4) Vaccinated (matched) COVID**_**pos**_: Number of COVID_pos_ cases among the vaccinated (matched) cohort, along with the percentage in parentheses, **(5) Unvaccinated (matched) COVID**_**pos**_: Number of COVID_pos_ cases among the unvaccinated (matched) cohort, along with the percentage in parentheses, **(6) Relative risk (95% CI):** Relative risk of COVID_pos_ in the vaccinated (matched) cohort compared to the unvaccinated (matched) cohort, along with 95% confidence interval in parentheses, **(7) BH-adjusted p-value:** Benjamini-Hochberg-adjusted Fisher exact test p-value.

| Vaccine | Race/<br>Ethnicity | Total<br>matched<br>pairs | Vaccinated<br>(matched)<br>COVID<sub>pos</sub> | Unvaccinated<br>(matched)<br>COVID<sub>pos</sub> | Relative risk<br>(95% CI) | BH-adjusted<br>p-value |
| --- | --- | --- | --- | --- | --- | --- |
| HepA-HepB | White | 5345 | 102 (1.91%) | 186 (3.48%) | 0.55 (0.43, 0.70) | 2.6E-05 |
| POLIO | White | 2182 | 32 (1.47%) | 74 (3.39%) | 0.43 (0.29, 0.66) | 9.9E-04 |
| MMR | White | 2032 | 44 (2.17%) | 86 (4.23%) | 0.51 (0.36, 0.73) | 3.3E-03 |
| Pneumococcal<br>conjugate (PCV13) | White | 5518 | 91 (1.65%) | 147 (2.66%) | 0.62 (0.48, 0.80) | 3.3E-03 |
| HIB | White | 1813 | 20 (1.1%) | 48 (2.65%) | 0.42 (0.25, 0.71) | 7.2E-03 |
| Diphtheria (with P/T) | White | 15008 | 406 (2.71%) | 493 (3.28%) | 0.82 (0.72, 0.94) | 0.03 |
| Geriatric Flu<br>Vaccine (65+ Yrs) | Black | 167 | 5 (2.99%) | 19 (11.4%) | 0.26 (0.11, 0.71) | 0.03 |
| Pneumococcal<br>conjugate (PCV13) | Black | 239 | 8 (3.35%) | 24 (10%) | 0.33 (0.16, 0.74) | 0.03 |
| Geriatric Flu<br>Vaccine (65+ Yrs) | White | 9511 | 144 (1.51%) | 192 (2.02%) | 0.75 (0.61, 0.93) | 0.05 |
| RZV Zoster<br>(ZOSTAVAX,<br>SHINGRIX) | Black | 206 | 8 (3.88%) | 22 (10.7%) | 0.36 (0.18, 0.81) | 0.06 |
| Meningococcal | Black | 133 | 39 (29.3%) | 22 (16.5%) | 1.77 (1.11, 2.78) | 0.08 |

**Table 9:** Summary of SARS-CoV-2 rates for race/ethnicity-stratified vaccinated and unvaccinated propensity score matched cohorts (5 year time horizon). Table of SARS-CoV-2 infection rates for vaccinated and unvaccinated (matched) race/ethnicity subgroup cohorts for vaccines administered within 2 years prior to PCR testing. Only rows with adjusted p-values ≤ 0.1 are included. Rows in which the SARS-CoV-2 rate is lower (adjusted p-value < 0.05) in the vaccinated cohort are highlighted in **green**, and rows in which the SARS-CoV-2 rate is lower in the unvaccinated cohort are highlighted in **orange**. The columns are **(1) Vaccine**: Name of the vaccine, **(2) Race/ethnicity**: Race/ethnicity subgroup, **(3) Total matched pairs**: Number of pairs from the propensity matching procedure, which is the sample size of both vaccinated and unvaccinated cohorts after matching, **(4) Vaccinated (matched) COVID**_**pos**_: Number of COVID_pos_ cases among the vaccinated (matched) cohort, along with the percentage in parentheses, **(5) Unvaccinated (matched) COVID**_**pos**_: Number of COVID_pos_ cases among the unvaccinated (matched) cohort, along with the percentage in parentheses, **(6) Relative risk (95% CI):** Relative risk of COVID_pos_ in the vaccinated (matched) cohort compared to the unvaccinated (matched) cohort, along with 95% confidence interval in parentheses, **(7) BH-adjusted p-value:** Benjamini-Hochberg-adjusted Fisher exact test p-value.

| Vaccine | Race/<br>Ethnicity | Total<br>matched<br>pairs | Vaccinated<br>(matched)<br>COVID<sub>pos</sub> | Unvaccinated<br>(matched)<br>COVID<sub>pos</sub> | Relative risk<br>(95% CI) | BH-adjusted<br>p-value |
| --- | --- | --- | --- | --- | --- | --- |
| Meningococcal | White | 5772 | 335 (5.8%) | 202 (3.5%) | 1.66 (1.40, 1.96) | 2.9E-07 |
| Pneumococcal<br>conjugate (PCV13) | White | 23706 | 413 (1.74%) | 581 (2.45%) | 0.71 (0.63, 0.81) | 2.0E-06 |
| POLIO | White | 3321 | 66 (1.99%) | 142 (4.28%) | 0.46 (0.35, 0.62) | 2.0E-06 |
| Meningococcal | Black | 460 | 123 (26.7%) | 61 (13.3%) | 2.02 (1.52, 2.65) | 6.4E-06 |
| Pneumococcal<br>conjugate (PCV13) | Black | 653 | 41 (6.28%) | 91 (13.9%) | 0.45 (0.32, 0.64) | 6.9E-05 |
| MMR | White | 5285 | 130 (2.46%) | 201 (3.8%) | 0.65 (0.52, 0.80) | 9.2E-04 |
| RZV Zoster<br>(ZOSTAVAX,<br>SHINGRIX) | Black | 359 | 15 (4.18%) | 44 (12.3%) | 0.34 (0.20, 0.61) | 9.7E-04 |
| HIB | Black | 170 | 10 (5.88%) | 31 (18.2%) | 0.32 (0.17, 0.65) | 5.5E-03 |
| Pediatric Flu<br>Vaccine | Black | 517 | 100 (19.3%) | 62 (12%) | 1.61 (1.20, 2.15) | 0.01 |
| TYPHOID | Black | 268 | 71 (26.5%) | 42 (15.7%) | 1.69 (1.20, 2.36) | 0.02 |
| Diphtheria (with P/T) | White | 38816 | 1153 (2.97%) | 1298 (3.34%) | 0.89 (0.82, 0.96) | 0.02 |
| VARICELLA | White | 4456 | 117 (2.63%) | 163 (3.66%) | 0.72 (0.57, 0.91) | 0.03 |
| HIB | White | 3731 | 62 (1.66%) | 97 (2.6%) | 0.64 (0.47, 0.88) | 0.03 |
| HepA-HepB | White | 12999 | 356 (2.74%) | 432 (3.32%) | 0.82 (0.72, 0.95) | 0.03 |
| Geriatric Flu Vaccine (65+ Yrs) | Black | 312 | 19 (6.09%) | 39 (12.5%) | 0.49 (0.29, 0.83) | 0.03 |
| HPV | Black | 354 | 84 (23.7%) | 56 (15.8%) | 1.50 (1.10, 2.02) | 0.04 |
| Pediatric Flu Vaccine | Asian | 463 | 35 (7.56%) | 17 (3.67%) | 2.06 (1.16, 3.54) | 0.05 |
| Geriatric Flu Vaccine (65+ Yrs) | White | 14410 | 226 (1.57%) | 281 (1.95%) | 0.80 (0.68, 0.96) | 0.05 |
| Influenza (any) | Black | 1181 | 171 (14.5%) | 132 (11.2%) | 1.30 (1.05, 1.60) | 0.06 |
| Pneumococcal conjugate (PCV13) | Hispanic | 897 | 66 (7.36%) | 93 (10.4%) | 0.71 (0.53, 0.96) | 0.10 |

We observe similar results for the colon cancer screening covariate. For example, the SARS-CoV-2 infection rate is 2.5% among persons with colon cancer screens in the past 5 years and 4.4% among persons without colon cancer screens in the past 5 years (p-value: 9.3e-44). After propensity score matching, the SARS-CoV-2 infection rate is 2.5% with and 2.4% without colon cancer screens in the past 5 years (p-value: 1). In total, 6 comparisons (2 controls, 3 time horizons each) were done. After applying Fisher’s method to combine p-values, we get a combined p-value of 0.22 (*X*^2^ = 15, df=12) against the combined hypothesis that none of the controls have an association with SARS-CoV-2 after propensity score matching.

We expect that the individuals who have recently taken cancer screens may have lower rates of SARS-CoV-2 infection due to the “healthy user effect”^17^. In particular, persons who have recently had mammograms or colonoscopies may engage in general health-seeking behaviors which decrease their risk of SARS-CoV-2 infection or generally decrease their risk of a positive PCR test result. The results from the negative control experiment demonstrates that the propensity score matching is able to correct for confounding variables which may contribute to spurious findings such as those caused by the healthy user effect.

### Tipping point analysis

In order to evaluate how robust the associations between vaccinations and SARS-CoV-2 infection found in this study are to the effects of potential confounders, we conduct a “tipping point” analysis^24^. The purpose of this analysis is to find the point at which an unobserved confounder would “tip” the conclusion on each vaccine, making the results no longer statistically significant.

Here, there are two dimensions to consider: (1) the effect size (i.e. relative risk of SARS-CoV-2 infection) of the confounder, and (2) the relative prevalence of the confounder in the vaccinated vs. unvaccinated (matched) cohorts. For each vaccine, we compute the relative prevalence and effect size that would be required for an unobserved confounder to overturn the conclusion for a given (vaccine, time horizon) pair. We present the results from the tipping point analysis in **Figure 6**.

### Institutional Review Board (IRB)

This research was conducted under IRB 20-003278, “Study of COVID-19 patient characteristics with augmented curation of Electronic Health Records (EHR) to inform strategic and operational decisions”. All analysis of EHRs was performed in a privacy-preserving environment. nference and the Mayo Clinic subscribe to the basic ethical principles underlying the conduct of research involving human subjects as set forth in the Belmont Report and strictly ensure compliance with the Common Rule in the Code of Federal Regulations (45 CFR 46) on the Protection of Human Subjects.

## Data Availability

The data analyzed was accessed under the IRB information provided herewith (IRB 20-003278), Study of COVID-19 patient characteristics with augmented curation of Electronic Health Records (EHR) to inform strategic and operational decisions (at the Mayo Clinic).

## Acknowledgements

The authors thank Murali Aravamudan, Patrick Lenehan, Walter Kremers, and Hilal Maradit-Kremers for their review and helpful feedback on this manuscript.

## Supplementary Material

**Figure S1:**
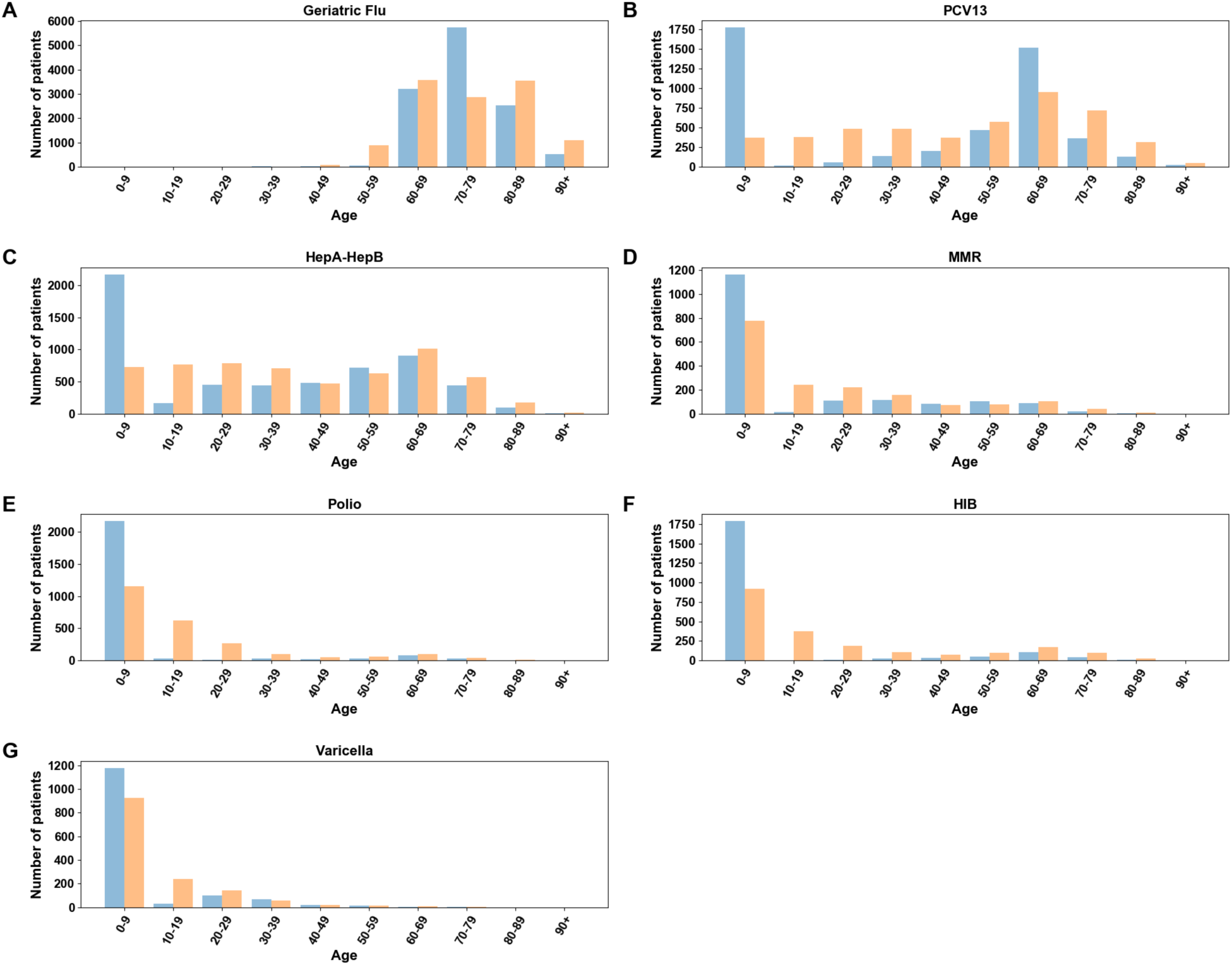
Age distributions in vaccinated (matched) and unvaccinated (matched) cohorts. For each vaccine associated with lower SARS-CoV-2 rates, age distributions for vaccinated (matched) and unvaccinated (matched) cohorts at the 1 year time horizon are shown. Vaccinated cohorts are shown in blue and unvaccinated cohorts are shown in orange. Numbers of patients in age ranges 0-9, 10-19, 20-29, 30-39, 40-49, 50-59, 60-69, 70-70, 80-80, and 90+ are shown for the following vaccines at the 1 year time horizon: **(A)** Geriatric Flu vaccine, **(B)** Pneumococcal Conjugate (PCV13), **(C)** Hepatitis A / Hepatitis B (HepA-HepB), **(D)** Measles-Mumps-Rubella (MMR), **(E)** Polio, **(F)** Haemophilus Influenzae type B (HIB), and **(G)** Varicella.

**Table S1.** Covariate balance for Geriatric Flu vaccine over 1-year time horizon. Mean/proportion values are shown for a selection of covariates for the vaccinated (matched), unvaccinated (matched), vaccinated (original), and unvaccinated (original) cohorts.

| **Covariate** | **Mean/**<br>**proportion**<br>**among**<br>**vaccinated**<br>**(n=12085)** | **Mean/proportion**<br>**among unvaccinated**<br>**(n=12085)** | **Unmatched**<br>**mean/proportion**<br>**among vaccinated**<br>**(n=13724)** | **Unmatched**<br>**mean/proportion among**<br>**unvaccinated**<br>**(n=123313)** |
| --- | --- | --- | --- | --- |
| **COVIDpos rate** | **1.57%** | **2.13%** | 1.49% | 4.44% |
| County incidence | 0.12% | 0.13% | 0.12% | 0.14% |
| County PCR test positive rate | 4.48% | 4.74% | 4.45% | 5.20% |
| **Age (std. dev)** | **74.8 (7.7)** | **74.5 (11.4)** | 75.4 (7.9) | 47.0 (21.1) |
| Gender - Male | 5823 (48.2%) | 5855 (48.4%) | 6632 (48.3%) | 54080 (43.9%) |
| Race - White | 11590 (95.9%) | 11502 (95.2%) | 13173 (96%) | 106806 (86.6%) |
| Race - Black | 158 (1.31%) | 203 (1.68%) | 171 (1.25%) | 5302 (4.3%) |
| Race - Asian | 127 (1.05%) | 133 (1.1%) | 154 (1.12%) | 3113 (2.52%) |
| Ethnicity - Hispanic | 185 (1.53%) | 211 (1.75%) | 198 (1.44%) | 7522 (6.1%) |
| Elixhauser - Hypertension | 9216 (76.3%) | 9041 (74.8%) | 10672 (77.8%) | 37095 (30.1%) |
| Elixhauser - Pulmonary | 4414 (36.5%) | 4171 (34.5%) | 5125 (37.3%) | 25010 (20.3%) |
| Elixhauser - Diabetes mellitus | 989 (8.18%) | 981 (8.12%) | 1121 (8.17%) | 5486 (4.45%) |
| Elixhauser - Diabetes mellitus (complications) | 2580 (21.3%) | 2409 (19.9%) | 3014 (22%) | 8787 (7.13%) |
| Elixhauser - Coagulopathy | 1687 (14%) | 1650 (13.7%) | 1968 (14.3%) | 7166 (5.81%) |
| Elixhauser - Obesity | 4404 (36.4%) | 4181 (34.6%) | 5077 (37%) | 28257 (22.9%) |
| Pregnancy - 90 days preceding | 3 (0.0248%) | 2 (0.0165%) | 4 (0.0291%) | 2558 (2.07%) |
| # unique other vaccines taken over preceding 5y | 3.93 | 3.63 | 4.05 | 1.65 |
| Propensity score | 0.82 | 0.80 | 0.84 | 0.16 |

**Table S2.** Covariate balance for Pneumococcal conjugate (PCV13) over 1-year time horizon. Mean/proportion values are shown for a selection of covariates for the vaccinated (matched), unvaccinated (matched), vaccinated (original), and unvaccinated (original) cohorts.

| Covariate | Mean/<br>proportion<br>among<br>vaccinated<br>(n=4693) | Mean/proportion<br>among unvaccinated<br>(n=4693) | Unmatched<br>mean/proportion<br>among vaccinated<br>(n=4693) | Unmatched<br>mean/proportion among<br>unvaccinated<br>(n=132344) |
| --- | --- | --- | --- | --- |
| **COVID<sub>pos</sub> rate** | **2.17%** | **3.03%** | 2.17% | 4.21% |
| County incidence | 0.12% | 0.12% | 0.12% | 0.14% |
| County PCR test<br>positive rate | 4.64% | 4.64% | 4.64% | 5.14% |
| **Age (std. dev)** | **38.7 (31.2)** | **48.8 (24.4)** | 38.7 (31.2) | 50.2 (21.4) |
| Gender - Male | 2404 (51.2%) | 2297 (48.9%) | 2404 (51.2%) | 58308 (44.1%) |
| Race - White | 4116 (87.7%) | 4192 (89.3%) | 4116 (87.7%) | 115863 (87.5%) |
| Race - Black | 197 (4.2%) | 170 (3.62%) | 197 (4.2%) | 5276 (3.99%) |
| Race - Asian | 108 (2.3%) | 102 (2.17%) | 108 (2.3%) | 3159 (2.39%) |
| Ethnicity - Hispanic | 301 (6.41%) | 222 (4.73%) | 301 (6.41%) | 7419 (5.61%) |
| Elixhauser -<br>Hypertension | 1728 (36.8%) | 2237 (47.7%) | 1728 (36.8%) | 46039 (34.8%) |
| Elixhauser -<br>Pulmonary | 1013 (21.6%) | 1390 (29.6%) | 1013 (21.6%) | 29122 (22%) |
| Elixhauser - Diabetes<br>mellitus | 200 (4.26%) | 278 (5.92%) | 200 (4.26%) | 6407 (4.84%) |
| Elixhauser - Diabetes<br>mellitus<br>(complications) | 539 (11.5%) | 737 (15.7%) | 539 (11.5%) | 11262 (8.51%) |
| Elixhauser -<br>Coagulopathy | 534 (11.4%) | 527 (11.2%) | 534 (11.4%) | 8600 (6.5%) |
| Elixhauser - Obesity | 1036 (22.1%) | 1404 (29.9%) | 1036 (22.1%) | 32298 (24.4%) |
| Pregnancy - 90 days preceding | 6 (0.128%) | 7 (0.149%) | 6 (0.128%) | 2556 (1.93%) |
| # unique other vaccines taken over preceding 5y | 4.37 | 4.74 | 4.37 | 1.79 |
| Propensity score | 0.66 | 0.66 | 0.66 | 0.34 |

**Table S3.**
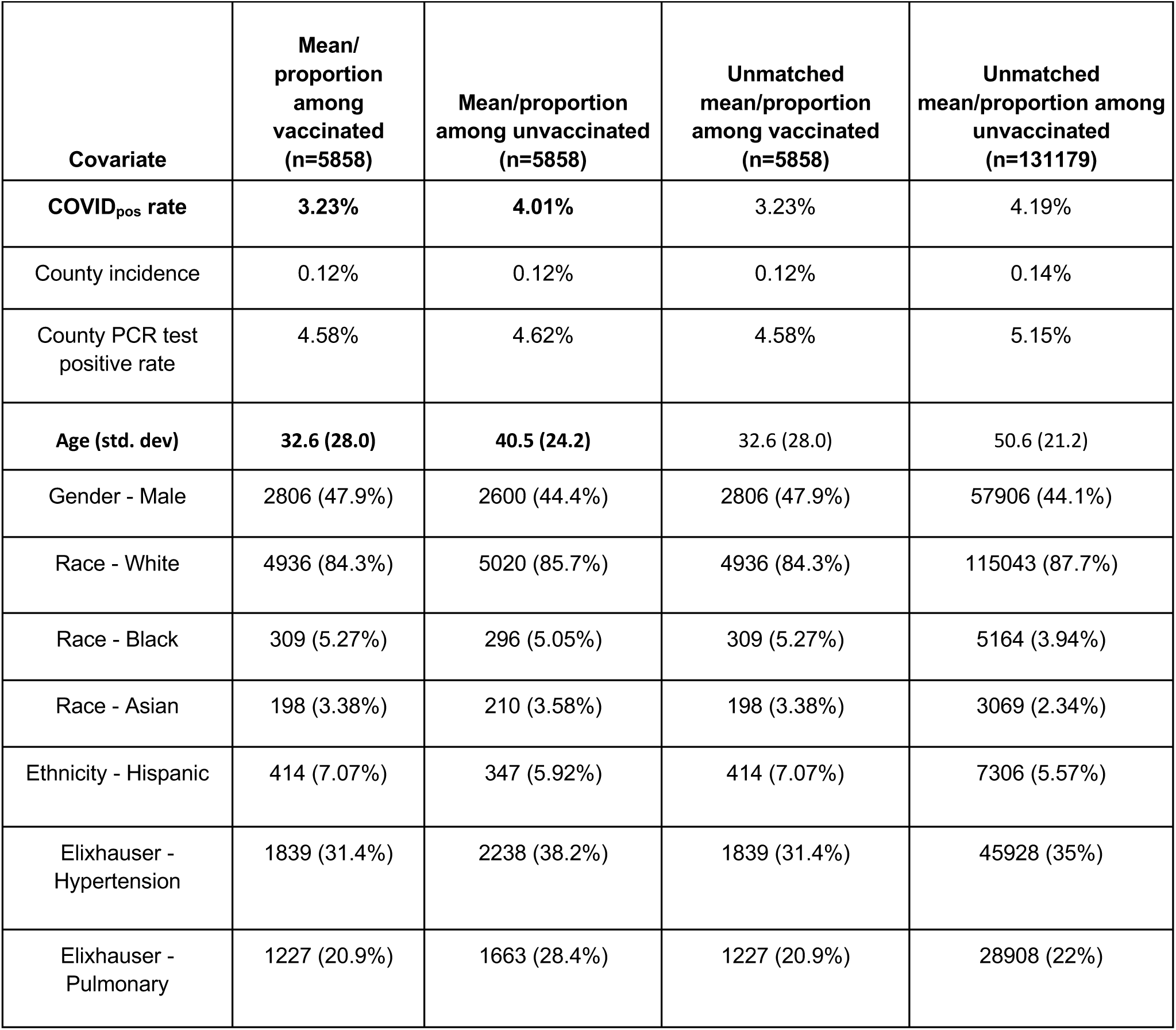
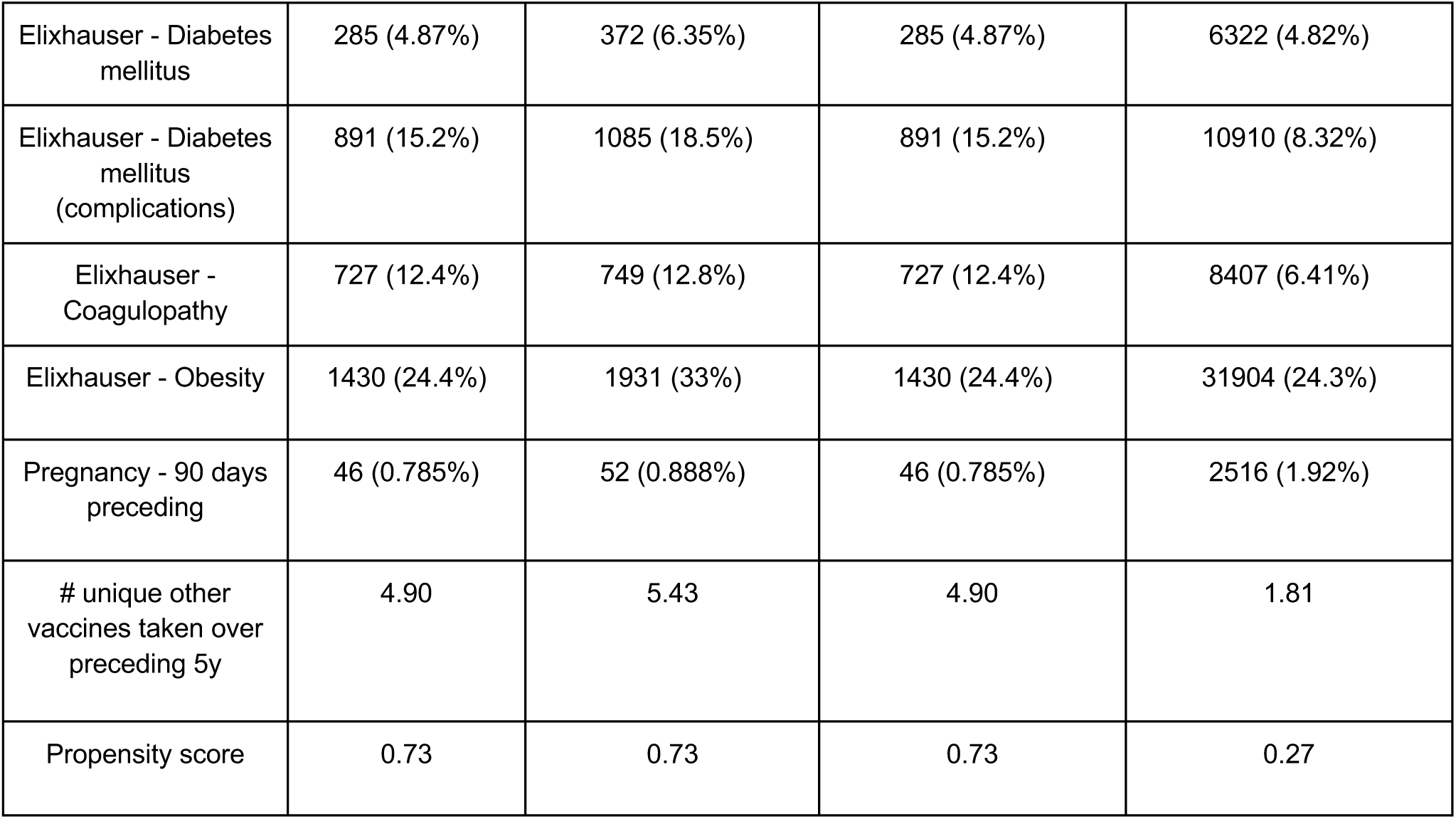
Covariate balance for HepA-HepB over 1-year time horizon. Mean/proportion values are shown for a selection of covariates for the vaccinated (matched), unvaccinated (matched), vaccinated (original), and unvaccinated (original) cohorts.

| Covariate | Mean/<br>proportion<br>among<br>vaccinated<br>(n=5858) | Mean/proportion<br>among unvaccinated<br>(n=5858) | Unmatched<br>mean/proportion<br>among vaccinated<br>(n=5858) | Unmatched<br>mean/proportion among<br>unvaccinated<br>(n=131179) |
| --- | --- | --- | --- | --- |
| **COVID<sub>pos</sub> rate** | **3.23%** | **4.01%** | 3.23% | 4.19% |
| County incidence | 0.12% | 0.12% | 0.12% | 0.14% |
| County PCR test positive rate | 4.58% | 4.62% | 4.58% | 5.15% |
| **Age (std. dev)** | **32.6 (28.0)** | **40.5 (24.2)** | 32.6 (28.0) | 50.6 (21.2) |
| Gender - Male | 2806 (47.9%) | 2600 (44.4%) | 2806 (47.9%) | 57906 (44.1%) |
| Race - White | 4936 (84.3%) | 5020 (85.7%) | 4936 (84.3%) | 115043 (87.7%) |
| Race - Black | 309 (5.27%) | 296 (5.05%) | 309 (5.27%) | 5164 (3.94%) |
| Race - Asian | 198 (3.38%) | 210 (3.58%) | 198 (3.38%) | 3069 (2.34%) |
| Ethnicity - Hispanic | 414 (7.07%) | 347 (5.92%) | 414 (7.07%) | 7306 (5.57%) |
| Elixhauser - Hypertension | 1839 (31.4%) | 2238 (38.2%) | 1839 (31.4%) | 45928 (35%) |
| Elixhauser - Pulmonary | 1227 (20.9%) | 1663 (28.4%) | 1227 (20.9%) | 28908 (22%) |
| Elixhauser - Diabetes mellitus | 285 (4.87%) | 372 (6.35%) | 285 (4.87%) | 6322 (4.82%) |
| Elixhauser - Diabetes mellitus (complications) | 891 (15.2%) | 1085 (18.5%) | 891 (15.2%) | 10910 (8.32%) |
| Elixhauser - Coagulopathy | 727 (12.4%) | 749 (12.8%) | 727 (12.4%) | 8407 (6.41%) |
| Elixhauser - Obesity | 1430 (24.4%) | 1931 (33%) | 1430 (24.4%) | 31904 (24.3%) |
| Pregnancy - 90 days preceding | 46 (0.785%) | 52 (0.888%) | 46 (0.785%) | 2516 (1.92%) |
| # unique other vaccines taken over preceding 5y | 4.90 | 5.43 | 4.90 | 1.81 |
| Propensity score | 0.73 | 0.73 | 0.73 | 0.27 |

**Table S4.**
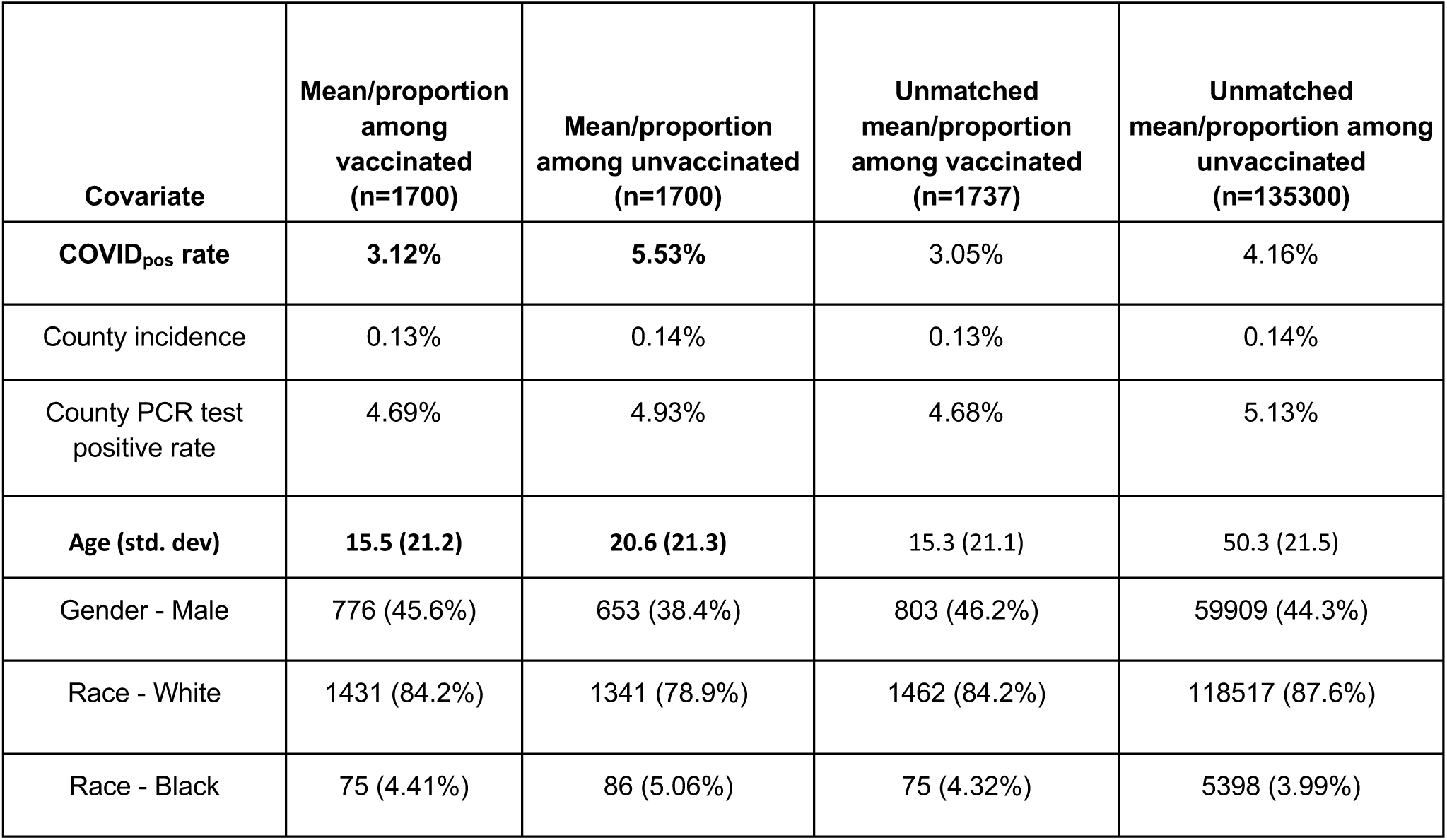
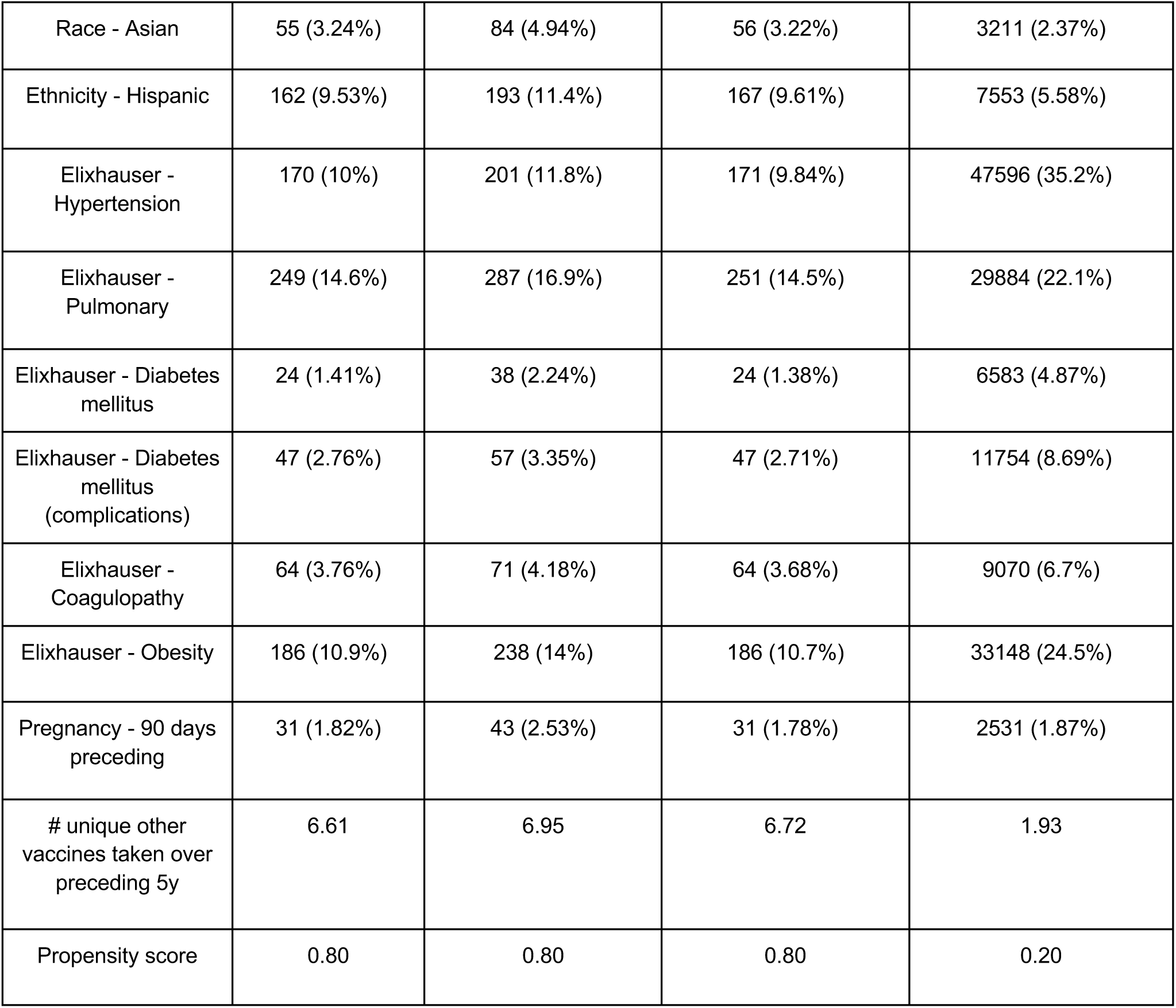
Covariate balance for MMR over 1-year time horizon. Mean/proportion values are shown for a selection of covariates for the vaccinated (matched), unvaccinated (matched), vaccinated (original), and unvaccinated (original) cohorts.

| Covariate | Mean/proportion among vaccinated (n=1700) | Mean/proportion among unvaccinated (n=1700) | Unmatched mean/proportion among vaccinated (n=1737) | Unmatched mean/proportion among unvaccinated (n=135300) |
| --- | --- | --- | --- | --- |
| **COVID<sub>pos</sub> rate** | **3.12%** | **5.53%** | 3.05% | 4.16% |
| County incidence | 0.13% | 0.14% | 0.13% | 0.14% |
| County PCR test positive rate | 4.69% | 4.93% | 4.68% | 5.13% |
| **Age (std. dev)** | **15.5 (21.2)** | **20.6 (21.3)** | 15.3 (21.1) | 50.3 (21.5) |
| Gender - Male | 776 (45.6%) | 653 (38.4%) | 803 (46.2%) | 59909 (44.3%) |
| Race - White | 1431 (84.2%) | 1341 (78.9%) | 1462 (84.2%) | 118517 (87.6%) |
| Race - Black | 75 (4.41%) | 86 (5.06%) | 75 (4.32%) | 5398 (3.99%) |
| Race - Asian | 55 (3.24%) | 84 (4.94%) | 56 (3.22%) | 3211 (2.37%) |
| Ethnicity - Hispanic | 162 (9.53%) | 193 (11.4%) | 167 (9.61%) | 7553 (5.58%) |
| Elixhauser - Hypertension | 170 (10%) | 201 (11.8%) | 171 (9.84%) | 47596 (35.2%) |
| Elixhauser - Pulmonary | 249 (14.6%) | 287 (16.9%) | 251 (14.5%) | 29884 (22.1%) |
| Elixhauser - Diabetes mellitus | 24 (1.41%) | 38 (2.24%) | 24 (1.38%) | 6583 (4.87%) |
| Elixhauser - Diabetes mellitus (complications) | 47 (2.76%) | 57 (3.35%) | 47 (2.71%) | 11754 (8.69%) |
| Elixhauser - Coagulopathy | 64 (3.76%) | 71 (4.18%) | 64 (3.68%) | 9070 (6.7%) |
| Elixhauser - Obesity | 186 (10.9%) | 238 (14%) | 186 (10.7%) | 33148 (24.5%) |
| Pregnancy - 90 days preceding | 31 (1.82%) | 43 (2.53%) | 31 (1.78%) | 2531 (1.87%) |
| # unique other vaccines taken over preceding 5y | 6.61 | 6.95 | 6.72 | 1.93 |
| Propensity score | 0.80 | 0.80 | 0.80 | 0.20 |

**Table S5.**
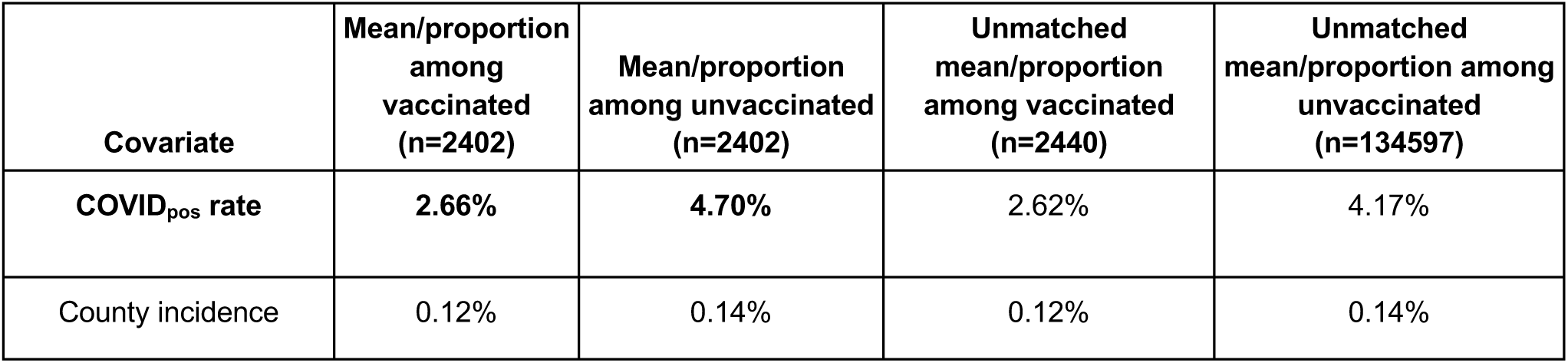
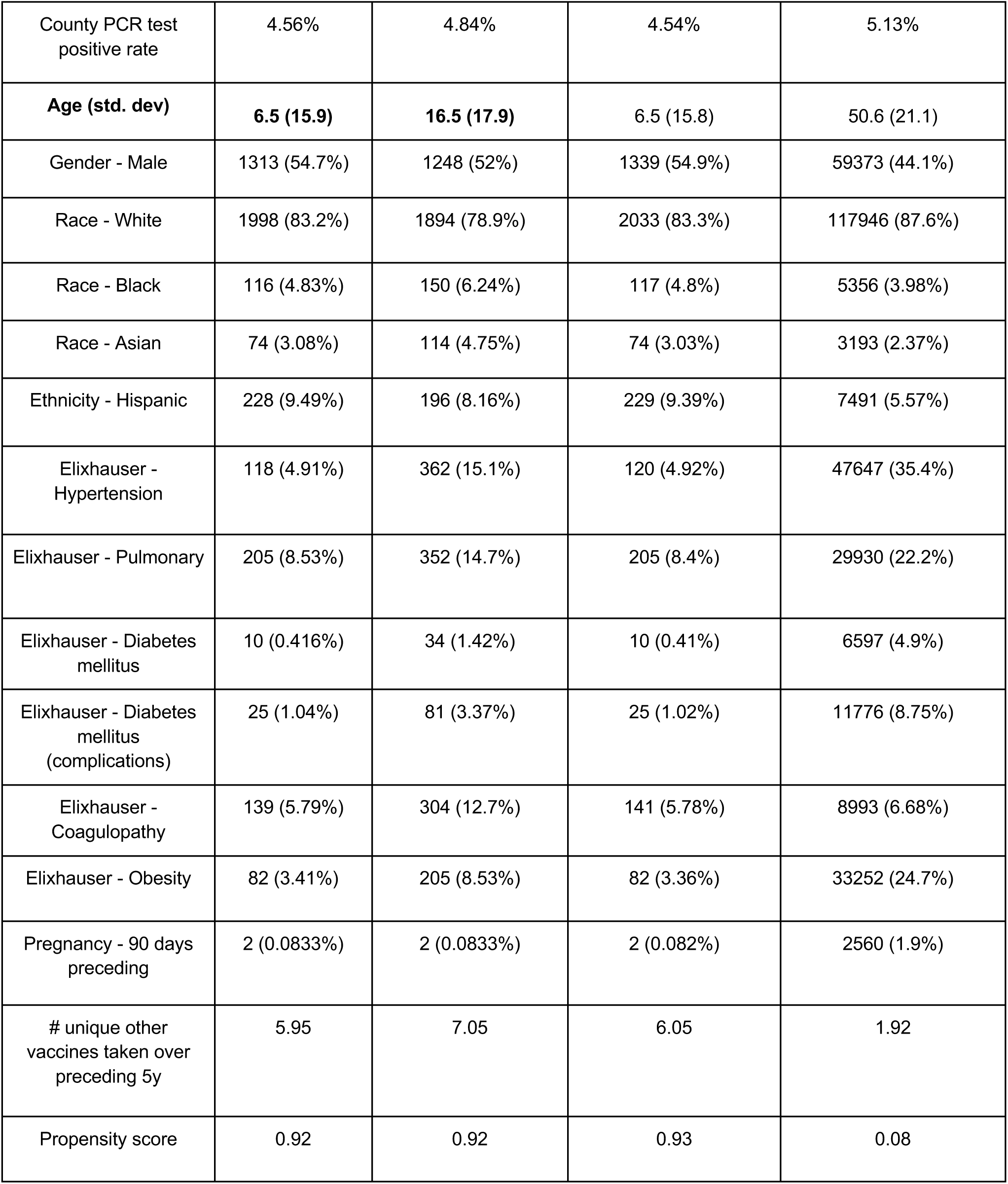
Covariate balance for Polio over 1-year time horizon. Mean/proportion values are shown for a selection of covariates for the vaccinated (matched), unvaccinated (matched), vaccinated (original), and unvaccinated (original) cohorts.

**Table S6.** Covariate balance for HIB over 1-year time horizon. Mean/proportion values are shown for a selection of covariates for the vaccinated (matched), unvaccinated (matched), vaccinated (original), and unvaccinated (original) cohorts.

| **Covariate** | **Mean/proportion among vaccinated (n=2061)** | **Mean/proportion among unvaccinated (n=2061)** | **Unmatched mean/proportion among vaccinated (n=2063)** | **Unmatched mean/proportion among unvaccinated (n=134974)** |
| --- | --- | --- | --- | --- |
| **COVID<sub>pos</sub> rate** | **2.09%** | **3.93%** | 2.08% | 4.18% |
| County incidence | 0.11% | 0.13% | 0.11% | 0.14% |
| County PCR test positive rate | 4.44% | 4.85% | 4.44% | 5.13% |
| **Age (std. dev)** | **8.7 (20.4)** | **22.8 (23.6)** | 8.6 (20.4) | 50.5 (21.3) |
| Gender - Male | 1120 (54.3%) | 1056 (51.2%) | 1121 (54.3%) | 59591 (44.1%) |
| Race - White | 1743 (84.6%) | 1753 (85.1%) | 1744 (84.5%) | 118235 (87.6%) |
| Race - Black | 87 (4.22%) | 78 (3.78%) | 87 (4.22%) | 5386 (3.99%) |
| Race - Asian | 57 (2.77%) | 66 (3.2%) | 57 (2.76%) | 3210 (2.38%) |
| Ethnicity - Hispanic | 188 (9.12%) | 175 (8.49%) | 188 (9.11%) | 7532 (5.58%) |
| Elixhauser - Hypertension | 176 (8.54%) | 512 (24.8%) | 178 (8.63%) | 47589 (35.3%) |
| Elixhauser - Pulmonary | 168 (8.15%) | 386 (18.7%) | 168 (8.14%) | 29967 (22.2%) |
| Elixhauser - Diabetes mellitus | 17 (0.825%) | 65 (3.15%) | 17 (0.824%) | 6590 (4.88%) |
| Elixhauser - Diabetes mellitus (complications) | 53 (2.57%) | 188 (9.12%) | 53 (2.57%) | 11748 (8.7%) |
| Elixhauser - Coagulopathy | 168 (8.15%) | 362 (17.6%) | 170 (8.24%) | 8964 (6.64%) |
| Elixhauser - Obesity | 85 (4.12%) | 285 (13.8%) | 86 (4.17%) | 33248 (24.6%) |
| Pregnancy - 90 days preceding | 1 (0.0485%) | 1 (0.0485%) | 1 (0.0485%) | 2561 (1.9%) |
| # unique other vaccines taken over preceding 5y | 5.10 | 6.50 | 5.11 | 1.95 |
| Propensity score | 0.90 | 0.89 | 0.90 | 0.11 |

**Table S7.** Covariate balance for Varicella over 1-year time horizon. Mean/proportion values are shown for a selection of covariates for the vaccinated (matched), unvaccinated (matched), vaccinated (original), and unvaccinated (original) cohorts.

| Covariate | Mean/<br>proportion<br>among<br>vaccinated<br>(n=1416) | Mean/proportion<br>among unvaccinated<br>(n=1416) | Unmatched<br>mean/proportion<br>among vaccinated<br>(n=1458) | Unmatched<br>mean/proportion among<br>unvaccinated<br>(n=135579) |
| --- | --- | --- | --- | --- |
| **COVID<sub>pos</sub> rate** | **2.75%** | **4.45%** | 2.88% | 4.16% |
| County incidence | 0.13% | 0.14% | 0.12% | 0.14% |
| County PCR test positive rate | 4.59% | 4.98% | 4.58% | 5.13% |
| **Age (std. dev)** | **7.3 (11.9)** | **10.2 (12.1)** | 7.2 (11.7) | 50.3 (21.5) |
| Gender - Male | 696 (49.2%) | 544 (38.4%) | 699 (47.9%) | 60013 (44.3%) |
| Race - White | 1203 (85%) | 1087 (76.8%) | 1239 (85%) | 118740 (87.6%) |
| Race - Black | 54 (3.81%) | 69 (4.87%) | 54 (3.7%) | 5419 (4%) |
| Race - Asian | 48 (3.39%) | 97 (6.85%) | 51 (3.5%) | 3216 (2.37%) |
| Ethnicity - Hispanic | 137 (9.68%) | 176 (12.4%) | 141 (9.67%) | 7579 (5.59%) |
| Elixhauser - Hypertension | 37 (2.61%) | 32 (2.26%) | 37 (2.54%) | 47730 (35.2%) |
| Elixhauser - Pulmonary | 159 (11.2%) | 146 (10.3%) | 161 (11%) | 29974 (22.1%) |
| Elixhauser - Diabetes mellitus | 6 (0.424%) | 8 (0.565%) | 6 (0.412%) | 6601 (4.87%) |
| Elixhauser - Diabetes mellitus (complications) | 10 (0.706%) | 12 (0.847%) | 10 (0.686%) | 11791 (8.7%) |
| Elixhauser - Coagulopathy | 38 (2.68%) | 43 (3.04%) | 38 (2.61%) | 9096 (6.71%) |
| Elixhauser - Obesity | 76 (5.37%) | 124 (8.76%) | 77 (5.28%) | 33257 (24.5%) |
| Pregnancy - 90 days preceding | 12 (0.847%) | 21 (1.48%) | 12 (0.823%) | 2550 (1.88%) |
| # unique other vaccines taken over preceding 5y | 6.98 | 7.12 | 7.12 | 1.94 |
| Propensity score | 0.88 | 0.88 | 0.89 | 0.11 |

